# Dietary preference and susceptibility to type 2 diabetes mellitus: a wide-angle Mendelian randomization study

**DOI:** 10.1101/2024.05.05.24306877

**Authors:** Mia D. Lee, Benjamin F. Voight

## Abstract

**Background:** Susceptibility to type 2 diabetes mellitus (T2D) is driven by genetic and environmental risk factors. Dietary preferences are a modifiable and largely environmental risk factor for T2D. The role of diet in disease liability has been limited to observational and epidemiologic studies with mixed findings.

**Objective:** To clarify the role of diet on susceptibility to T2D using genetic variants associated dietary preferences.

**Methods:** We collected genome-wide association data for 38 dietary preference traits plus T2D and 21 related cardiometabolic traits. We performed Mendelian randomization (MR) using genetic variants to test causal hypotheses between diet as the exposure and T2D or cardiometabolic traits as outcomes using univariable and multivariable methods along with the MR Robust Adjusted Profile Score (MR-RAPS) approach to increase power. We performed mediation analyses to evaluate the effects of dietary preferences on T2D to elucidate potential causal graphs and estimate the effects of dietary preferences mediated by potential mediators.

**Results:** We report 17 significant relationships between dietary preferences and T2D or a cardiometabolic risk factor (Bonferroni-corrected P < 5.99 x 10^-5^), including that higher intake of cheese, dried fruit, muesli, or fat-based spreads protected against T2D. We detected 7 additional associations (Bonferroni-corrected P < 1 x 10^-4^), with inclusion of additional genetic variants in MR-RAPS analysis. In multivariable MR, we discovered that body mass index (BMI) was a common, shared mediator for many of these observed associations. In mediation analysis, we confirmed that substantial proportions of the protective effects of cheese, dried fruit and muesli intakes on T2D were mediated by BMI. We further observed that educational attainment was an additional mediator exclusively for muesli intake-T2D association.

**Conclusions:** Our results provide genetic evidence supporting a link between diet and body weight, and are in line with observation of obesity and T2D in individuals and their specific preferences for food.

## INTRODUCTION

Type 2 diabetes mellitus (T2D) is a complex metabolic disease with individual risk influenced by both genetic and environmental factors [1,2]. The growing incidence of T2D in parallel with the obesity epidemic suggests the essential role of environment to disease risk [3]. While genome-wide association studies (GWAS) have elucidated the polygenic nature of the disease with discovery of hundreds of associated genetic loci [4,5], the spectrum of the environmental components that cause T2D is poorly understood. Improvements in this understanding could facilitate personalized interventions for patients who are at the most risk of cardiovascular, neurological and renal complications of diabetes based on these factors [6].

Despite observational data supporting the role of diet as a major but modifiable risk factor associated with T2D [7,8], a causal role of dietary factors and cardiometabolic disease is far less established with only a handful of examples in evidence [9,10]. Previous work supporting strong association between unhealthy diet and T2D has been largely accrued through observational studies in which a formal assessment of causality is difficult if not impossible. Moreover, previous reports on the effects of diet on susceptibility to T2D have been inconsistent and inconclusive [11-13], compounding the challenge of establishing a robust assessment of causality. Thus, a key challenge is to elucidate how specific dietary risk factors – or potentially how established risk factors like obesity – mediate or cause diabetes, and subsequently how to use these insights to action downstream interventional studies.

While non-genetic contributions to dietary preference are large and central, some food intake patterns have been observed to be heritable [14,15] and interact with T2D-relevant genes [16], suggesting that genetics may contribute a small component to interindividual variability in dietary preferences. Recently, GWAS have reported genetic associations with macronutrient intake, dietary habits, and food preferences [17-19]. Excitingly, these data offer the opportunity to examine evidence of causal relationships between diet and cardiometabolic outcomes using statistical approaches.

In this report, we utilized the framework of Mendelian randomization (MR) using genetic summary data related to diet and 42 dietary preferences to understand the role of these factors for T2D susceptibility and other cardiometabolic traits (**Figure 1**). MR is one statistical approach designed to infer causal effects between an exposure and an outcome. This method leverages genetic variants associated with an exposure (here, dietary preference) as instrumental variables (IVs) to estimate the effect of that exposure on an outcome (e.g., liability to T2D) [20]. MR has been used to confirm or refute a range of risk factors for T2D [22-23], including other diseases [24-26], as well as using dietary instrumental variables as exposures [27]. We developed instruments for 42 dietary preferences as exposures and performed univariable and multivariable experiments using T2D and 21 cardiometabolic risk factors as outcomes.

**Figure 1:**
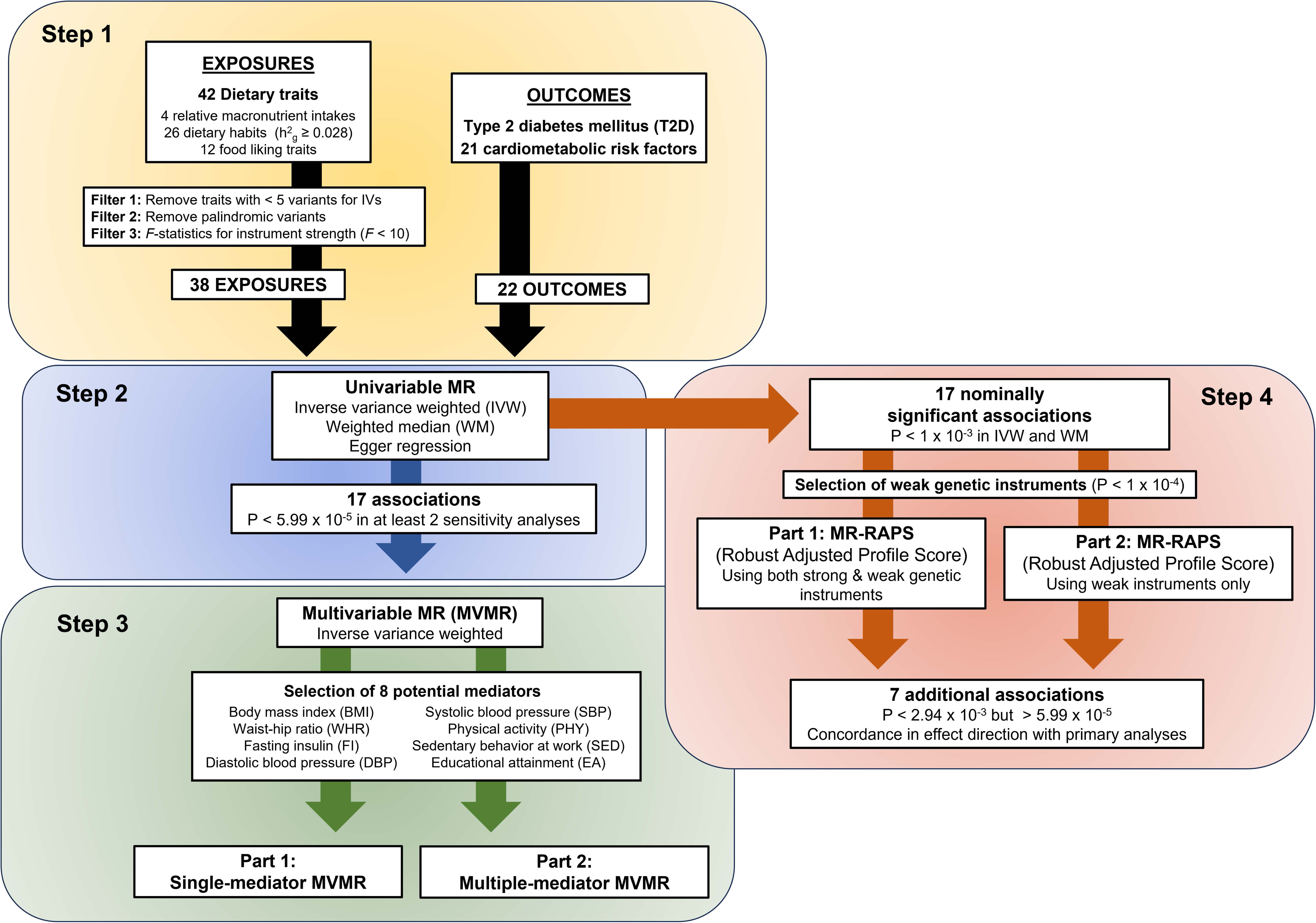
Schematic workflow of the Mendelian Randomization study on diet and T2D. This figure describes how the MR study on diet and T2D was conducted in 4 steps to investigate their relationship and evaluate the causality. After selecting viable genetic variants that meet the given criteria, univariable MR analysis estimated the total effects of dietary preferences on T2D and 21 related cardiometabolic risk factors. Then, MR-RAPS, which is an alternative method to standard MR, included weak instruments to re-evaluate the nominally significant exposure:outcome associations and detect additional associations. Finally, the significant associations from UVMR and MR-RAPS were further assessed by estimating the direct effects of dietary preferences on the outcomes after accounting for potential mediators in MVMR. Abbreviations – MR, Mendelian Randomization; MR-RAPS, Mendelian Randomization Robust Adjusted Profile Score; T2D, Type 2 diabetes mellitus; UVMR, Univariable Mendelian randomization.

## METHODS

### Acquisition of dietary trait summary genetic data

For dietary traits, we selected 42 dietary preferences, which were composed of: (i) 4 relative macronutrient intake measures, (ii) 26 dietary habits, and (iii) 12 food-liking traits. The summary statistic association data for these dietary traits were obtained from 3 studies on diet composition [17], dietary habits [18] and food liking [19] (**Supplementary Table 1**). These studies were conducted mainly on groups of individuals similar to the 1000 Genomes Project Continental group from Europe as a reference.

Based on 24h dietary food recall questionnaire, which contains ≥70 food items assessed in UK Biobank participants, the summary association data for diet composition, which was adjusted for sex, year of birth, and the first 10 genetic principal components, identified genetic associations with relative intake of carbohydrate, sugar, fat and protein [17]. The carbohydrate intake included a consumption of all saccharides, while the sugar intake mono- and disaccharides only. The measurements for 4 macronutrient intakes were corrected for total energy intake. The energy values of the macronutrients were obtained using the conversion factor of 4 kcal/gram for protein, sugar and carbohydrate, and 9 kcal/gram for fat.

The summary association data for dietary habits, in which covariates were age in months, sex and 10 genetic principal components, was based on UK Biobank food frequency questionnaire (FFQ) on 85 single food items on daily and/or weekly basis [18]. The measurement of the intake of each food item was in either quantitative continuous variables (e.g., cups per day, pieces per day, bowls per week) or ordinal non-quantitative variables (e.g., number of intakes per day or week). The GWAS for dietary habits also estimated SNP heritability (h^2^) of those single food intake using BOLT-lmm software (v.2.3.2) pseudo-heritability measurement representing the fraction of phenotypic variance explained by the estimated relatedness matrix [18]. We selected 26 specific dietary habits that were in the top 25 percentile of the observed SNP heritability (h^2^ > 0.028).

The summary association data obtained for food liking conducted an assessment for likings for 139 specific foods and beverages using the food preferences/liking questionnaire. This questionnaire uses the 9-point hedonic scale, where 1 corresponds to “Extremely dislike” and 9 to “Extremely like”. The GWAS for food liking included age, sex and the first 10 genetic principal components as covariates for the analysis. Because these foods and beverages are not generalized and overlap with some of the dietary habits, we selected 3 main dimensions and 9 sub-dimensions of food likings that were categorized by taste and food type in the multi-level hierarchical manner [19].

### Acquisition of T2D and related traits summary genetic data

For T2D, we obtained the latest summary statistics from Million Veteran Program (MVP) study [4], which included age, sex and ten genetic principal components as covariates. We selected the following 21 cardiometabolic risk factors related to T2D: Body mass index (BMI) [28], Waist-hip ratio (WHR) [28], Waist-hip ratio adjusted for BMI (WHRadjBMI) [28], Fasting glucose adjusted for BMI (FG) [29], Fasting insulin adjusted for BMI (FI) [29], Glycated hemoglobin (HbA1c) [29], High-density lipoprotein cholesterol (HDL) [30], Low-density lipoprotein cholesterol (LDL) [30], Triglyceride (TG) [30], Abdominal subcutaneous adipose tissue volume (ASAT) [31], Visceral adipose tissue volume (VAT) [31], Pancreas fat (Pancfat) [34128465], Pancreas iron content (Panciron) [31], Pancreas volume (Pancvol) [31], Liver fat (Liverfat) [31], Liver iron content (Liveriron) [31], Liver volume (Livervol) [31], Alkaline phosphatase (ALP) [32], Alanine aminotransferase (ALT) [32], Aspartate transaminase (AST) [33], and Gamma-glutamyl Transferase (GGT) [32]. For reasons of statistical power, we focused primarily on European ancestry data, which had the largest sample sizes. The locations, sample sizes and other information for these summary data are detailed in **Supplementary Table 2**.

### GWAS data processing and quality control

Prior to selection of genetic instruments for dietary traits, we first removed rare variants (MAF < 0.01) and, if applicable, variants with low imputation quality (INFO < 0.5) in the summary-level data sets for dietary traits (**Supplementary Figure 2**). As additional quality control, we also eliminated duplicate variants. Finally, we obtained consensus rsIDs for all variants (dbSNP build 155) and aligned them to GRCh37 for all traits (see **Code Availability**).

### Selection of variants for genetic instruments for dietary traits

We used PLINK (v1.9 software) [34] and European 1000 Genomes Project Phase 3 data (EUR, GRCh37) for LD reference to group loci together using the ‘LD clumping’ procedure to select genome-wide significant (P < 5 x 10^-8^) and independent genetic instruments (instrumental variables) with low pairwise linkage with one another (r^2^ < 0.001) within a window of 500kb [21,35] (Supp. Figure 2). After clumping, we excluded four dietary traits from the MR analysis because they had relatively few IVs contributing (less than n = 5): Flora spread intake, low-fat spread intake, low-fat milk intake and liking for healthy foods.

We removed palindromic (A/T or C/G) genetic variants that are present in the dietary traits to ensure robustness of our instruments (**Supplementary Table 3).** The genetic variants that were present in the dietary traits but not the outcomes of interest (e.g., T2D and related traits) were removed from MR analysis and not replaced with proxy variants. With this approach, we identified 931 genetic instruments (803 unique SNPs) in total across 38 dietary traits. The final genetic instruments used for MR are shown in the **Supplementary Tables 14 and 15**, and the SNP selection process is described in **Supplementary Figure 1**.

For each genetic instrument, we estimated R^2^, the proportion of variance explained of exposure by created IV, assuming a strictly additive model [36]. For a single SNP R^2^ given by: 2 × *p* × (1 – *p*) × β^2^, where *p* is the minor allele frequency and β is the estimated effect size of the SNP for the exposure trait [36]. The total variance explained then is simply the sum across all SNPs, R^2^_total_. Once we obtained R^2^_total_, for each exposure, we calculated the F-statistic (*F*) to assess the strength of the instrumental variables. The formula for *F* is (N – K – 1)/K × R^2^_total_ /(1 – R^2^_total_), where N is the sample size, and K is the number of SNPs used for the IV [37]. All of our generated instruments exceeded an *F* > 10, suggesting minimal effects from weak instrument bias (**Supplementary Table 4**).

We used the LD-clumping procedure described above using 1000 Genomes Project Phase 3 (EUR) as the LD reference to identify the best LD proxy (r^2^ ≥ 0.8 with given index variants) for index variants that were present with dietary genetic instruments but not present in the summary association trait data used in multivariable MR (MVMR) analyses.

For the analysis using MR-RAPS (Robust Adjusted Profile Score) [38], we used the same approach described above (LD-clumping via PLINK, [34]), and applied the same parameters to obtain independent, sub-genome wide (P < 1 x 10^-4^) significant genetic variants as additional instruments. As described above for the primary MR analyses, we removed palindromic genetic variants in the MR-RAPS analysis.

### Mendelian randomization methods

MR is a statistical causal inference analysis that employs genetic variants associated with an exposure (here, dietary preference) as instrumental variables (IVs) to estimate the effect of that exposure on an outcome (e.g., liability to T2D) [20]. Specific assumptions for instrumental variables are required, namely that: (i, Relevance assumption) genetic variants are associated with the exposure, (ii, Independence assumption) genetic variants are not associated with any potential confounders in the exposure-outcome association, and that (iii, Exclusion restriction assumption) genetic variants can influence the outcome only through the exposure (Supplementary Figure 1). Given Mendel’s law of segregation and independent assortment at meiosis, the design of an MR experiment is analogous to the design of the randomized control trial via random allocation of genetic variants [21] and can address issues of confounding and reverse causality in assessment of causality.

We populated STROBE-MR checklist as a guideline to conduct all our MR analyses and report the results clearly (**Supplementary Data 1**).

For primary analysis, we performed two-sample univariable MR (UVMR) using summary statistic association data from 38 dietary traits as exposures and from T2D plus 21 T2D-related cardiometabolic traits as outcomes. Though partial sample overlap exists between dietary traits and some outcome traits, the bias due to the observed sample overlap were estimated to be small in the analysis, especially when the strength of genetic instruments estimated by F-statistic (*F*) is strong (*F* > 10) and the datasets are of different sample size [39] (**Supplementary Table 21**). Using TwoSampleMR package (v.0.5.8) [40,41], we employed three standard UVMR methods to estimate causal effects: Inverse variance weighted under the random effects model (IVW), Weighted median (WM), and Egger regression. Each method has different sensitivity for the IV assumptions. IVW has the greatest statistical power because it assumes all variants do not violate any IV assumptions; WM assumes at least half of IVs are valid instruments, providing robustness to Assumptions 2 and 3; Egger regression estimates the causal effect after accounting for horizontal pleiotropy in IVs, providing robustness to Assumption 3 [42]. We used the Egger bias intercept test as a quantitative indicator of bias due to horizontal pleiotropy (**Supplementary Table 5**). After exclusion for instrument size (see above) and checking the presence of exposure IVs in the endpoint data, we performed UVMR that tested a total of 835 exposure-outcome pairwise associations (Supplementary Table 5). We also performed MR-Steiger test [41] to examine whether assumption that exposure causes outcome is valid and determine the directionality of those associations by using the same input data that were used to estimate effects via the 3 standard UVMR methods.

To address the burden of multiple testing, we considered Bonferroni correction to determine the adjusted nominal p-value threshold for significance (n=835 tests with a 5% error rate: P < 5.99 x 10^-5^). We considered Bonferroni-corrected exposure-outcome associations further if two additional criteria were met: (i) they were associated across at least two MR analyses, and (ii) at least 5 IVs were used in the sensitivity analyses.

### Analysis using MR-RAPS (Robust Adjusted Profile Score)

17 nominally but not multi-test corrected significant (i.e., MR P < 1 x 10^-3^ but P > 5.99 x 10^-5^) exposure-outcome associations across at least two sensitivity analyses were further analyzed using the MR-Robust Adjusted Profile Score (MR-RAPS) method. MR-RAPS minimizes measurement error in SNP-exposure effects due to weak instruments bias and is robust to systematic pleiotropy [38]. In addition to genome-wide significant variants (P < 5 x 10^-8^), sub-genome wide significant (P < 1 x 10^-4^) genetic variants as weak instruments were included to increase the power of MR for causal inference and re-assess those associations via this method.

For statistical stringency, we applied the following criteria to determine whether the associations are significant and additional associations were identified by increased power from MR-RAPS: (i) when estimated using the combined set of both weak (P < 1 x 10^-4^) and strong (P < 5 x 10^-8^) instruments, the association exceeds the threshold p-value that was applied for the primary UVMR analysis (e.g., P < 5.99 x 10^-5^); (ii) when estimated using only weak (P < 1 x 10^-4^) instruments, the association exceeds the threshold p-value that was applied for the primary UVMR analysis (P < 5.99 x 10^-5^); and (iii) the effect direction of the association that meets those two criteria above was concordant with that observed from the initial UVMR methods described above.

### Multivariable MR experimental methods

To further evaluate exposure-outcome associations that emerged as significant from UVMR, we used MVMR package (v.0.4) [43] along with the TwoSampleMR package (v.0.5.8) and performed multivariable MR (MVMR) using IVW to estimate the direct effects of dietary preferences by accounting for the indirect effects mediated through selected potential mediators (**Supplementary Figure 3**). Since T2D is a multi-factorial, complex disease, we relied on biological prior and literature-based observations in T2D to narrow the list of potential traits that could possibly mediate the causal effects of exposures on outcomes. Then, we conducted Linkage Disequilibrium Score Regression (LDSC) analysis [44] on those traits to ensure that they are genetically correlated with T2D and dietary exposures of interest (**Supplementary Tables 17-18**). Based on these factors, we selected 8 potential mediators: Body mass index (BMI) [28], Fasting insulin (FI) [29], Waist-hip ratio (WHR) [28], Diastolic blood pressure (DBP) [45], Systolic blood pressure (SBP) [45], Physical activity (PHY) [46] and Sedentary behavior at work (SED) [46], and Educational Attainment (EA, Supplementary Table 2) [47].

Our MVMR analysis consisted of two parts: single-mediator MVMR and multiple-mediator MVMR. For both single-mediator and multiple-mediator MVMR, the genetic instruments selected for potential mediators and outcomes were based on the genetic variants that were considered strong, viable genetic instruments for the primary exposures (here, dietary preferences), namely exposure-centric instruments. In other words, all the exposure-centric instruments were the same instruments utilized for UVMR.

In single-mediator MVMR, each of the 8 selected potential mediators was applied in a pairwise manner to estimate the direct effects of the primary exposures on T2D and related cardiometabolic traits as outcomes. In the multiple-mediator MVMR, at least 2 potential mediators were included when assessing the effect of the primary exposures on the outcomes.

### Mediation analysis

We employed a two-step MR approach to conduct mediation analysis, which facilitates estimation of the indirect effect of a primary exposure on an outcome via a mediating trait and, consequently, the proportion mediated. For this analysis, the two steps are: (i) estimation of the effect of a primary exposure on a mediating trait, and (ii) estimation of the direct effect of a mediating trait on an outcome, after adjusting for a primary exposure [48,49].

We applied this approach to 3 exposure-outcome pairs of our main interest which were associations of cheese, muesli and dried fruits intake with T2D. Spreads intake association with T2D was excluded due to a very small number of genetic instruments available for spreads intake. For mediation analysis, BMI, DBP and EA were the mediating traits of interest because they attenuated those 3 associations in single-mediator MVMR.

In the first step, we used “exposure-centric” instruments and obtained the effects of the 3 dietary preferences on BMI, DBP and EA individually via UVMR. In the second step, we estimated the direct effects of BMI, DBP and EA on T2D, accounting the effects of the dietary preferences, via MVMR. Because the 3 mediators were considered the primary exposures in the second step, we utilized all the genetic variants that serve as strong genetic instruments for each mediator, namely mediator-centric instruments, to obtain estimates of the second step with increased statistical power. We chose IVW as a main method for both steps.

We then employed product of coefficients method to multiply the estimates (i.e., the effect sizes in beta coefficients) from the two steps and obtained the indirect effects of the 3 dietary preferences on T2D. These indirect effects were divided by the total effects of the dietary preferences on T2D estimated in the primary UVMR to calculate the proportions mediated by BMI, DBP and EA, respectively.

Finally, we used RMediation package (v. 1.2.2) [50] to estimate indirect effects and approximate standard errors and 95% confidence intervals, using 3 different methods: Monte Carlo, distribution of product and asymptotic distribution (i.e., delta). The package uses the effect sizes in beta coefficients and standard errors of the two steps including a parameter *rho*, which is a correlation (-1 < *rho* < 1) between the two steps which are exposure-to-mediator and mediator-to-outcome pathways. We assumed the default value of 0 for this parameter for the 3 methods and they yielded similar confidence intervals for indirect effects. The indirect effect estimates from these 3 methods are comparable to the ones computed via product of coefficients method. Of the 3 methods, we relied on the estimates mainly from delta method, which is relatively more suitable for summary-level data in two-sample MR [51,52].

## RESULTS

### Univariable MR detected 17 associations of dietary preferences with T2D and cardiometabolic risk factors

To examine the relationship between diet and T2D, we first performed two-sample MR on 38 dietary traits as exposures and T2D plus 21 additional cardiometabolic risk factors as outcomes (**Methods,** Supplementary Tables 1-2). **Supplementary Table 3** shows the number of genetic instruments our for 38 dietary traits, and we did not observe evidence of potential weak instrument bias from our generated genetic IVs as calculated by *F*-statistics (**Supplementary Table 4**). Of the 835 exposure-outcome pairs we tested, we considered only those significant after Bonferroni multiple test correction (nominal P < 5.99 x 10^-5^, **Methods**). Of 835 associations, we excluded associations where less than 5 genetic instruments were used in the analyses. We applied three univariable MR (UVMR) statistics for our initial approach: Inverse variance weighted (IVW, random effects model), Weighted median (WM) and MR-Egger, requiring that at least two achieved a Bonferroni corrected p-value to claim significance (**Methods**). After filtering, we observed non-zero causal effect estimates for 17 exposure-outcome relationship (**Figure 2**). While 16 of them were robust specifically through IVW and WM (Supplementary Table 5), only one exceeded the given threshold p-value in all 3 sensitivity analyses.

**Figure 2:**
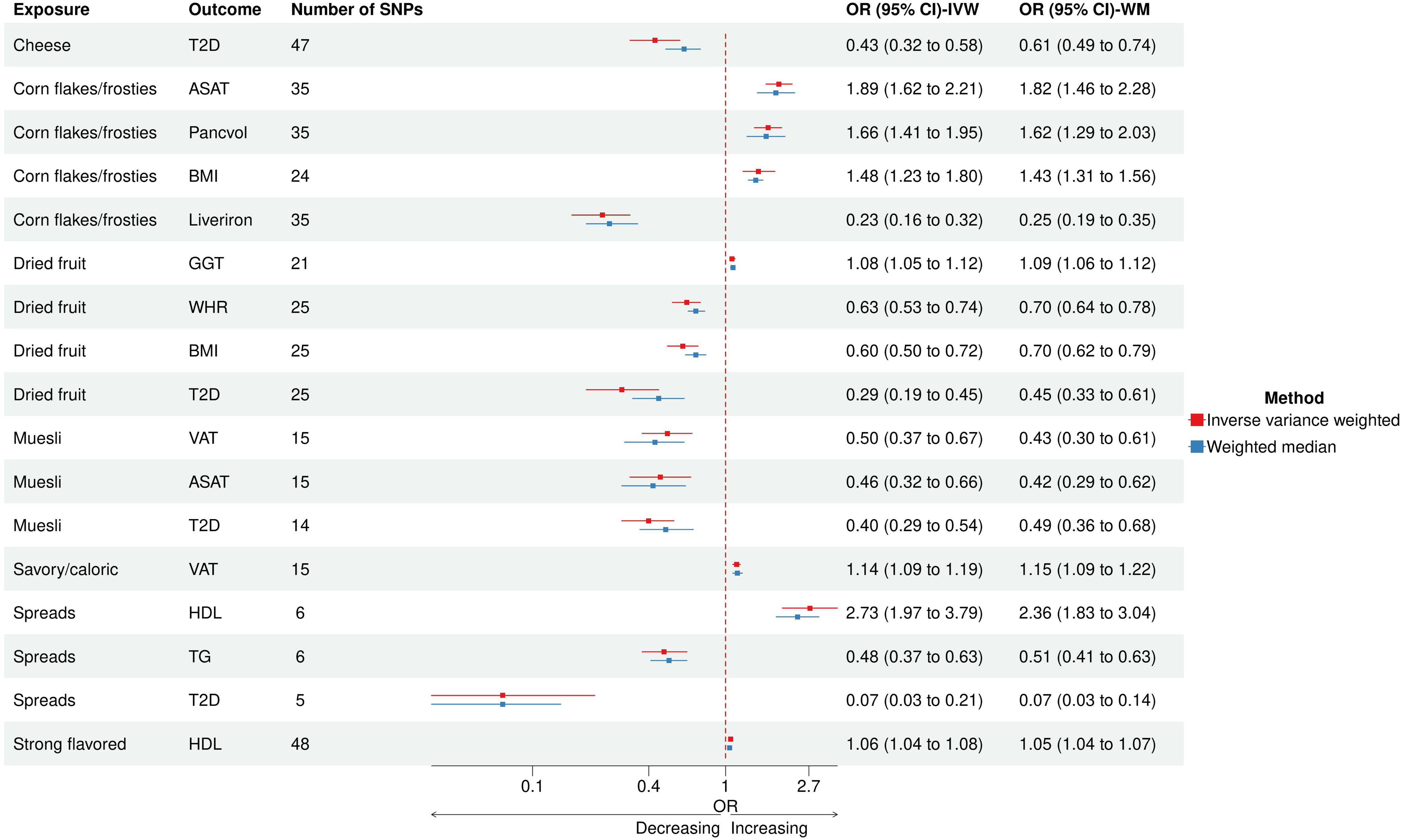
Forest plot for 17 associations from UVMR. This figure is a forest plot for 17 associations from UVMR-IVW and UVMR-WM in descending order of OR for each dietary preference trait. Given the Bonferroni-corrected p-value, these associations passed the significance threshold (P < 5.99 x 10-5) in MR using IVW and WM methods. Abbreviations – IVW, Inverse variance weighted; MR, Mendelian Randomization; OR, Odds ratio; UVMR, Univariable Mendelian randomization; WM, Weighted median.

We observed that four dietary preferences had inverse associations with T2D. A one-SD increment in cheese intake (CHEESE), which was measured in a number of consumptions per day and per week, lowered T2D risk by 57% (IVW, β = -0.84; OR, 0.43 per 1 SD; 95% confidence (CI), 0.32 to 0.58; P = 2.91x 10^-8^). A one-SD higher intake of dried fruit (DRIEDFRU), which was measured in a number of pieces per day, also lower T2D risk (IVW, β = -1.23; OR, 0.29; 95% CI, 0.19 to 0.45; P = 2.49 x 10^-8^). Intake of muesli (MUESLI), which is primarily composed of rolled oats, was also observed to reduce the risk of T2D (IVW, β = -0.93; OR, 0.40; 95% CI, 0.36 to 0.68; P = 2.58 x 10^-9^). As a type of cereal, muesli intake was measured in a number of bowls per week. Lastly, intake of spreads (SPREADS), which consist of number of uses of various fat-based spreads, was shown to have the largest reduction in T2D risk (IVW β = -2.60; OR, 0.07; 95% CI, 0.02 to 0.21; P = 1.41 x 10^-6^), though we note that only five genetic instruments were used to assess the effect of spread intake against T2D.

### Intake of dried fruits, muesli, spreads and corn flakes/frosties are causally associated with cardiometabolic risk factors

Consistent with the above effect of diet on T2D susceptibility, dried fruit, muesli and spread intake also were related to several cardiometabolic risk factors. The protective effect of dried fruit intake on T2D was reflected through lower body mass index (BMI) (IVW, β = -0.51; OR, 3.21; 95% CI, 0.60 to 0.72; P = 1.27 x 10^-8^) and lower waist-hip ratio (WHR) (IVW, β = -0.46; OR, 0.63; 95% CI, 0.53 to 0.74; P = 5.88 x 10^-8^). Also, dried fruit intake was shown to have a mild reducing effect on the liver function measure of gamma-glutamyl transferase (GGT) (IVW, β = -0.08; OR, 0.92; 95% CI, 0.90 to 0.95; P = 5.25 x 10^-7^). Likewise, muesli intake lowered volumes of two adiposity-relevant organs: abdominal subcutaneous adipose tissue (ASAT) (IVW, β = -0.77; OR, 0.46; 95% CI, 0.32 to 0.66; P=2.45 x 10^-5^) and visceral adipose tissue (VAT) (IVW, β = -0.70; OR, 0.50; 95% CI, 0.37 to 0.67; P = 2.64 x 10^-6^). Intake of spreads elevated high-density lipoprotein (HDL) (IVW β = 1.00; OR, 2.73; 95% CI, 1.97 to 3.79; P=1.91 x 10^-9^) and lower triglycerides (TG) (IVW β = -0.73; OR, 0.48; 95% CI, 0.37 to 0.63; P=1.17 x 10^-7^). However, we note that only six genetic instruments were used to assess the relationship between spreads intake and these two lipids.

In addition, intake of corn flakes/frosties (CORNFLAK), which was based on a number of bowls per week, increased ASAT by 89% (IVW, β = 0.64; OR, 1.89; 95% CI, 1.62 to .21; P = 6.55 x 10^-15^) and BMI by 49% (IVW, β = 0.40; OR, 1.49; 95% CI, 1.23 to 1.80; P = 4.65 x 10^-5^). Intake of Corn flakes/frosties also elevated Pancreas volume (Pancvol) (IVW, β = 0.50; OR, 1.66; 95% CI, 1.41 to 1.95; P=8.94 x 10^-10^) but lowered Liver iron content (Liveriron) (IVW, β = -1.48; OR, 0.23; 95% CI, 0.16 to 0.32; P = 1.70 x 10^-17^). Corn flakes/frosties intake to Liver iron content was the only association tested to be significant across all 3 sensitivity analyses. The MR-Egger causal estimate of this association was consistent with that of IVW and WM, and here the MR-Egger intercept was 0.04 (P = 2.28 x 10^-3^, Supplementary Table 5), suggesting a suppressive effect. Corn flakes and frosties are processed, refined carbohydrate-based foods with a high glycemic index. Hence, the associations of corn flakes/frosties intake with these two organs suggest the presence of dietary role in affecting the health of diabetes-relevant organs in the development of T2D. Together, the relationships between these four dietary traits and cardiometabolic risk factors agree with observational data that suggests both central adiposity and dyslipidemia are strongly linked to T2D susceptibility [53,54].

### Preferences for Savory/Caloric foods and Strong-flavored foods are causally linked with cardiometabolic risk factors

Finally, UVMR revealed two food preference traits related with cardiometabolic risk factors. Preference for savory/caloric (SAVCAL) food increased VAT (IVW, β = 0.13; OR, 1.14; 95% CI, 1.09 to 1.19; P = 2.70 x 10^-9^). SAVCAL falls under one of main food liking categories, namely ‘highly-palatable’, which consists of high caloric, energy-dense foods, such as meat and deep-fried foods. Preference for strong flavored (STR) food increased HDL (IVW, β = 0.06; OR, 1.06; 95% CI, 1.04 to 1.08; P = 7.42 x 10^-12^), though the effect size was relatively modest. STR is a part of the other main food liking category, ‘acquired’, which consists of bitter, spicy, sharp flavored foods, such as coffee and cheese, where preferences are acquired throughout life.

To evaluate the directionality of the causal effect estimated between dietary traits and outcomes, we conducted the MR-Steiger test (**Supplementary Table 6**) [41]. We observed that 14 of the 17 exposure-outcome associations were predicted to have the causal effect direction indicated by UVMR analyses. For the remaining three, the test showed significant reverse causality (corn flakes/frosties intake to Liver iron content, Steiger P = 3.23 x 10^-19^) or was inconclusive on this test (muesli intake to ASAT and VAT, Steiger P > 0.05). The possible explanations for the opposite direction of causality could be due to the presence of pleiotropy in genetic instruments and unmeasured confounding, which can be examined through inclusion of potential mediators in multivariable MR (MVMR).

### MR-RAPS included weak genetic instruments and detected 7 additional associations

To identify additional exposure-outcome associations that could provide more information about the Diet-T2D relationship, we employed MR-RAPS (Robust Adjusted Profile Score), an alternative to the 3 standard MR methods, to re-assess 17 nominally significant (P < 1 x 10^-3^) (**Supplementary Table 7**) associations in at least 2 sensitivity analyses. Thus, in addition to strong instruments selected for UVMR, we included weak instruments, which are sub-genome wide significant (P < 1 x 10^-4^), for MR-RAPS analysis.

Out of 17 associations, we identified seven additional exposure-outcome effects that met our criteria for significance (**Methods, Figure 3**). These associations surpassed the Bonferroni-adjusted p-value (P < 5.99 x 10-5) applied in the primary MR analysis (**Supplementary Table 8**) and were in the correct direction of causality based on Steiger’s test (**Supplementary Table 9**). The directionality of these seven associations estimated by MR-RAPS and standard MR methods was concordant, although as expected the effect size estimated from in MR-RAPs based on a larger panel of SNP (P < 1 x 10^-3^) was substantially lower in many cases, relative to instruments from genome-wide significant variants (P < 5 x 10^-8^).

**Figure 3:**
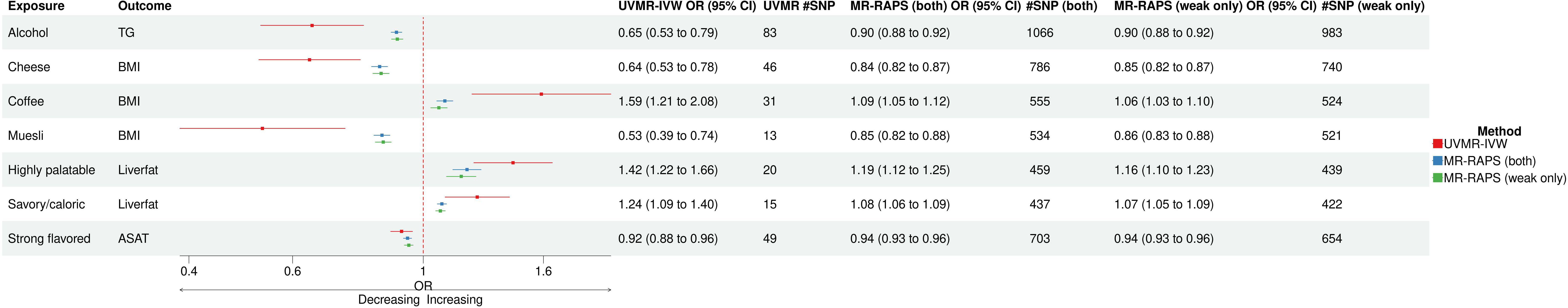
Forest plot for 7 associations from MR-RAPS. This figure is a forest plot for 7 associations identified via MR-RAPS. In comparison, the results from MR-RAPS analysis using only weak instruments and using both strong and weak instruments are consistent in direction with that of IVW method in UVMR. Abbreviations – IVW, Inverse variance weighted; MR-RAPS, Mendelian Randomization Robust Adjusted Profile Score; UVMR, Univariable Mendelian Randomization.

In MR-RAPS, elevated preference for cheese consumption lowered BMI (β = -0.18; OR, 0.84 per 1 SD; 95% CI, 0.82 to 0.86; P=1.35 x 10^-33^) and higher preference for muesli also lowered BMI (β = -0.16; OR, 0.85 per 1 SD; 95% CI, 0.83 to 0.88; P = 6.15x 10^-28^). These 2 associations were in line with the protective effects of cheese and muesli intake on T2D observed in the primary analysis. Also, they support that BMI is a potential mediator of the associations of these 2 dietary traits with T2D. Conversely, preference for coffee intake was shown to elevate BMI (β = 0.09; OR, 1.09 per 1 SD; 95% CI, 1.06 to 1.13; P = 1.19 x 10^-9^). Additionally, MR-RAPS suggested a negative link between alcohol intake and triglycerides (β = -0.11; OR, 0.90 per 1 SD; 95% CI, 0.88 to 0.92; P = 7.51 x 10^-28^).

Preference for STR was linked to slightly lower ASAT (β = -0.06; OR, 0.94 per 1 SD; 95% CI, 0.93 to 0.96; P= 3.30 x 10^-15^). Preference for SAVCAL, which was shown to have increasing effect on VAT in the primary analysis, elevated liver fat content (Liverfat) (β = 0.07; OR, 1.07 per 1 SD; 95% CI, 1.05 to 1.09; P = 7.11 x 10^-17^). Preference for highly palatable foods (PAL), whose sub-category is SAVCAL, increased liver fat content (β = 0.17; OR, 1.18 per 1 SD; 95% CI, 1.12 to 1.24; P = 5.91 x 10^-11^).

We also explored MR-RAPS with spreads intake and HDL, TG and T2D, given only 5-6 genetic instruments were utilized in UVMR. With inclusion of weak instruments, the relationship between spreads intake and these traits did not reject the null hypothesis, and the strong causal effects, particularly on HDL and TG, estimated in UVMR were no longer observed (**Supplementary Table 10**). Given the high p-value, the extremely large effect of spreads intake on T2D was greatly reduced with additional instruments (β = -0.083; OR, 0.92 per 1 SD; 95% CI, 0.82 to 1.03; P = 0.15).

### Multivariable MR suggests that BMI is a common mediating risk factor for dietary exposure-outcome relationships

To evaluate the 17 associations observed in the UVMR and better understand the underlying causal pathways on outcomes, we next conducted multivariable MR (MVMR) experiments. We employed 8 well-established non-dietary T2D risk factors as potential mediators (**Methods**), including BMI, WHR, Fasting insulin adjusted for BMI (FI), Educational attainment (EA), Diastolic blood pressure (DBP), Systolic blood pressure (SBP), Physical activity (PHY) and Sedentary behavior at work (SED, Supplementary Table 2). We found that the causal effects observed for 15 exposure-outcome associations were either substantially or fully attenuated after including BMI in single-mediator MVMR (**Figure 4, Supplementary Table 11**), which assessed the effect of an exposure on an outcome after adjustment for a single mediating trait in a pairwise manner. For example, BMI appeared to be responsible for mediating the protective effect of cheese intake against T2D, as no other trait attenuated this association (Supplementary Table 11). In addition, BMI completely attenuated the association between SAVCAL and VAT as well as the associations of corn flakes/frosties intake with ASAT, Liver iron content, and Pancreas volume. Furthermore, BMI yielded the largest attenuation in spreads intake to T2D and spreads intake to HDL associations. This observation shows that the preference of various spreads is closely linked with adiposity and therefore could contribute to cardiometabolic health.

**Figure 4:**
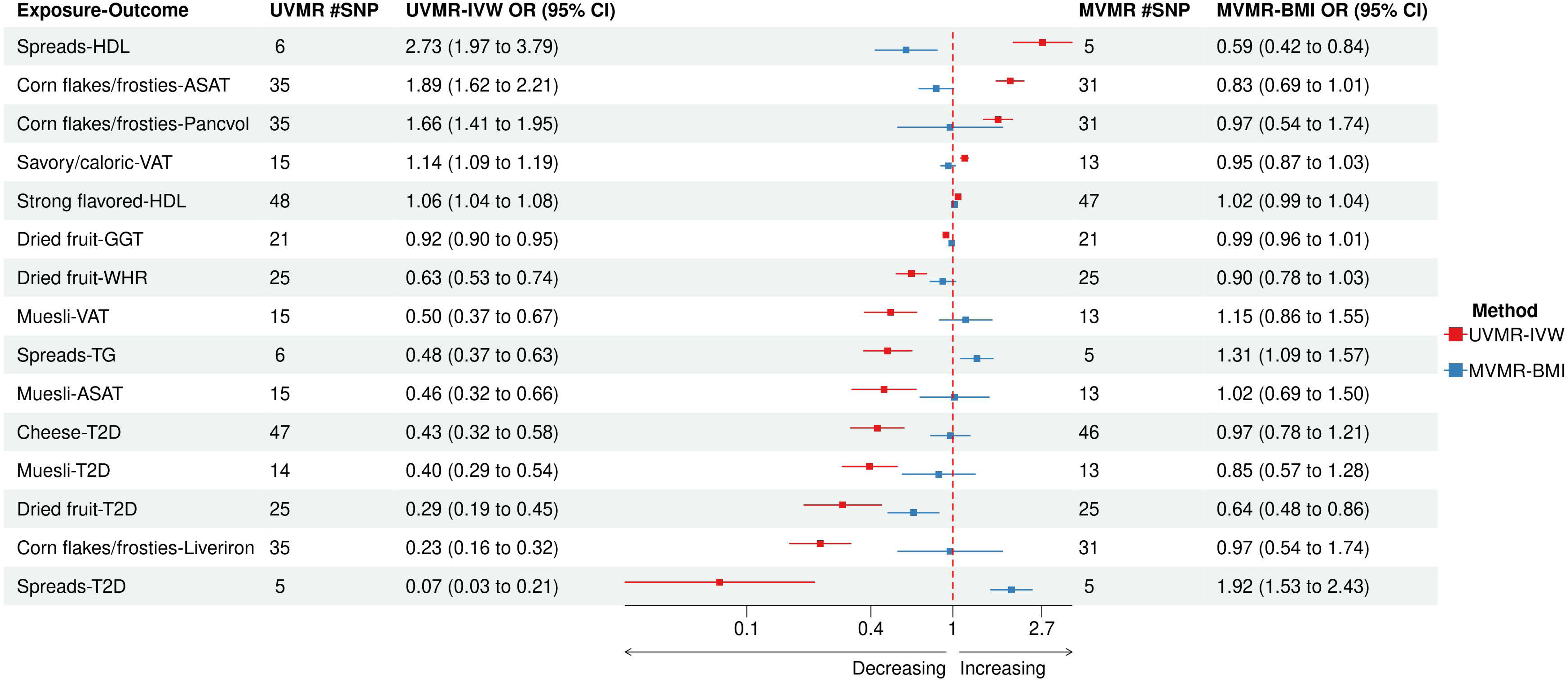
Forest plot for single-mediator MVMR using BMI vs. UVMR-IVW. This figure is a forest plot that displays the results of both single-mediator MVMR using BMI as a potential mediator and UVMR-IVW on 15 associations. Compared to the results of univariable MR using IVW, the strengths of 15 associations were either partially or completely attenuated by BMI. Abbreviations – BMI, Body mass index; IVW, Inverse variance weighted; MVMR, Multivariable Mendelian Randomization; UVMR, Univariable Mendelian Randomization.

Since the 4 dietary associations with T2D were our primary interest, we also checked whether there were additional factors that attenuated any of those associations beyond BMI (Supplementary Table 11). The single-mediator MVMR suggested that DBP could be an additional driver for the observed effect of dried fruit intake on BMI, GGT, T2D and WHR and the observed effect of muesli intake on ASAT, T2D and VAT. However, the effect of DBP as a potential mediator on these associations were not clearly determinable (P > 0.05) and segregated relatively wide 95% confidence intervals. Surprisingly, there was residual direct effect of dried fruit intake on T2D even after accounting the mediating effect of BMI in the single-mediator MVMR. We observed that EA was an additional factor that attenuated the associations of muesli intake with T2D, ASAT and VAT, to a degree that comparable to those by BMI (**Figure 5**).

**Figure 5:**
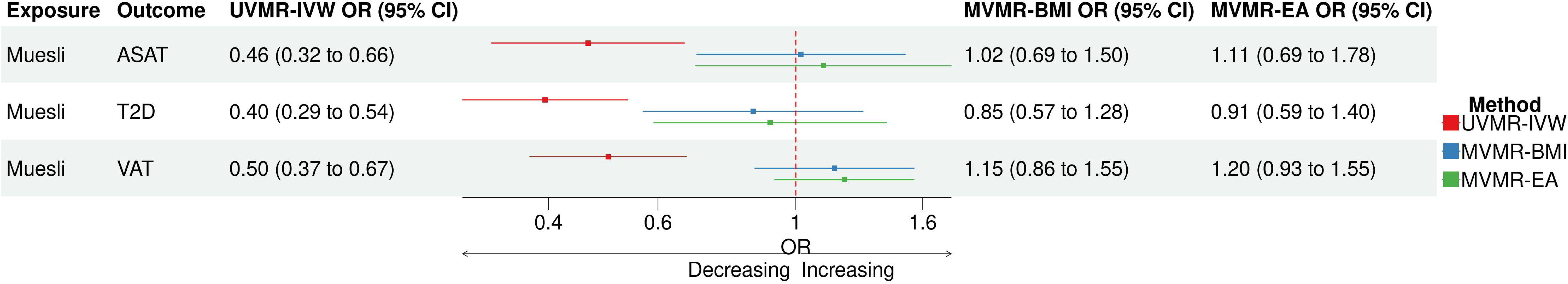
Forest plot for single-mediator MVMR using BMI and EA on Muesli intake associations. This figure is a forest plot for Muesli-ASAT, T2D and VAT associations in UVMR and single-mediator MVMR. Aside from BMI, EA was an additional factor that attenuated the muesli intake associations, to a degree that comparable to those by BMI. Abbreviations – ASAT, Abdominal subcutaneous adipose tissue; BMI, Body mass index; EA, Educational attainment; MVMR, Multivariable Mendelian Randomization; T2D; UVMR, Univariable Mendelian Randomization; VAT, Visceral adipose tissue.

Followed by single-mediator MVMR, we then conducted additional MVMR experiments to re-estimate the effect of the exposures in joint with multiple mediators on the outcomes. We carried out 2 different multiple-mediator MVMR analyses: non-BMI mediators MVMR and all-inclusive MVMR, which includes all selected potential mediating traits. The purpose of multiple-mediator MVMR was to verify whether BMI is the primary mediators of the associations amongst the selected potential mediating traits. In non-BMI mediators MVMR, we checked whether non-BMI mediators together as a group can yield greater attenuation in the absence of BMI. In all-inclusive MVMR, we examined the direct effect of the exposures on the outcomes by accounting the summative effect of all mediators and compared with that of non-BMI mediators MVMR.

Overall, the multiple-mediator MVMR showed that BMI was a major mediator for most of the associations observed (**Supplementary Figure 4, Supplementary Table 12**). For few associations, it was not immediately clear if BMI was the strongest mediator. The multiple-mediator MVMR further supported that BMI was a definitive primary risk factor for mediating the protective effect of cheese intake against T2D. These analyses also indicated that summative effect of multiple mediators does not necessarily result in greater attenuation.

### BMI was a common mediator for the 3 dietary associations identified by MR-RAPS

For further assessment, we performed single-mediator MVMR on the seven additional associations detected in MR-RAPS (**Supplementary Table 13**). The same eight potential mediators were employed to determine whether same, non-dietary risk factors mediate these associations. Notably, none of the potential mediators attenuated the Cheese Intake-BMI and ALC-TG associations. This underscores our observation that BMI is the primary mediator for the effect of cheese intake on T2D. BMI was shown to attenuate PAL-Liver fat content, SAVCAL-Liver fat content and STR-ASAT association (**Figure 6**). EA was also a mediator for muesli intake-BMI, SAVCAL-Liver fat content, and STR-ASAT associations (**Figure 7**). These two findings were congruent with the observations in the MVMR on the dietary associations from UVMR.

**Figures 6:**
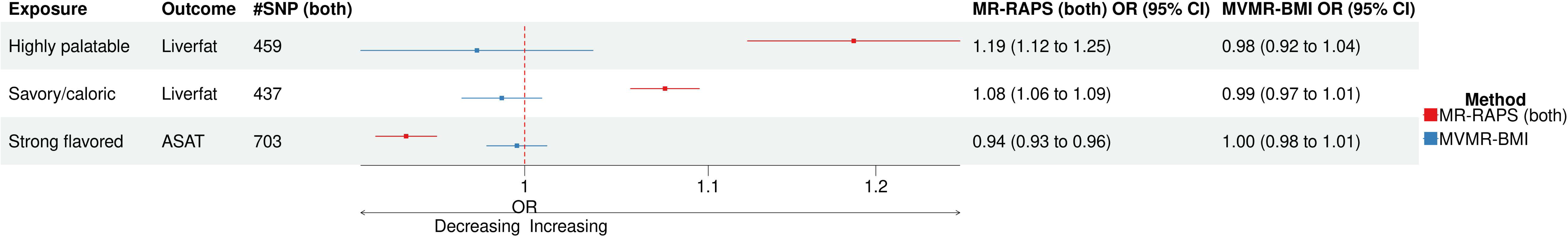
Forest plot for single-mediator MVMR using BMI on the associations from MR-RAPS. This figure is a forest plot for single-mediator MVMR using BMI on the following associations detected in MR-RAPS: PAL-Liver fat content, SAVCAL-Liver fat content and STR-ASAT associations. As observed in single-mediator MVMR on the associations in UVMR, BMI was a common mediating trait that attenuated the effects of these 3 dietary preferences on Liver fat content and ASAT. Abbreviations – ASAT, Abdominal subcutaneous adipose tissue; BMI, Body mass index; MR-RAPS, Mendelian Randomization Robust Adjusted Profile Score; MVMR, Multivariable Mendelian Randomization; PAL, Highly palatable; SAVCAL, Savory/ caloric; STR, Strong flavored; UVMR, Univariable Mendelian Randomization.

**Figures 7:**
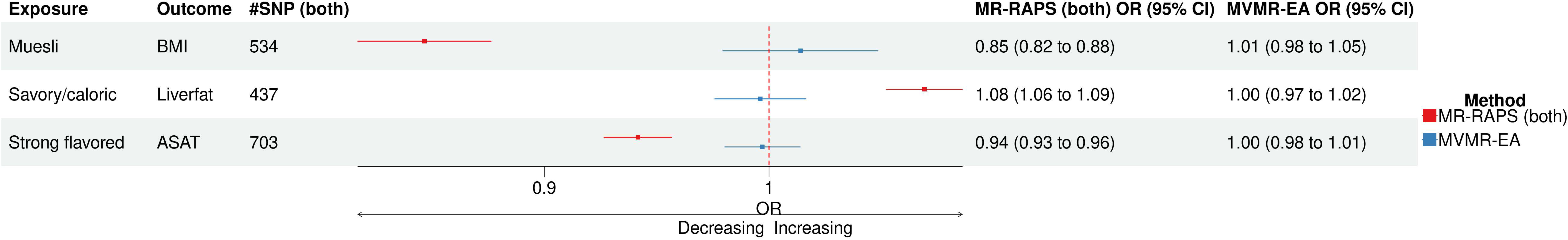
Forest plot for single-mediator using EA on the associations from MR-RAPS. This figure is a forest plot for single-mediator MVMR using EA on the following associations detected in MR-RAPS: SAVCAL-Liver fat content, STR-ASAT and muesli intake-BMI associations. Forest plot for single-mediator MVMR on 3 associations in MR-RAPS. Consistent with the observations in MVMR on the associations, particularly with muesli intake, in UVMR, EA was another mediating factor for dietary associations with cardiometabolic risk factors. Abbreviations – ASAT, Abdominal subcutaneous adipose tissue; BMI, Body mass index; EA, Educational attainment; MR-RAPS, Mendelian Randomization Robust Adjusted Profile Score; MVMR, Multivariable Mendelian Randomization; SAVCAL, Savory/ caloric; STR, Strong flavored.

### BMI was a major contributor to the dietary associations with T2D in the mediation analysis

In addition to MVMR, we took two-step MR approach (**Methods**) to conduct mediation analysis and estimate the indirect effects of cheese, dried fruit and muesli intake on T2D after accounting for BMI, DBP and EA individually as mediating traits (**Supplementary Tables 19-20**). Spreads intake-T2D association was excluded due to a small number of strong genetic instruments (n=5) used to test the association. Whereas MVMR estimates the direct effect of a primary exposure on outcome, mediation analysis quantifies the indirect effect mediated by a secondary exposure. We compared the results of mediation analysis to those of MVMR and evaluated those 3 mediating traits as potential mediators for the dietary associations with T2D. **Figure 8** displays the proportion of the effect of each dietary preference on T2D explained by each mediator separately.

**Figure 8:**
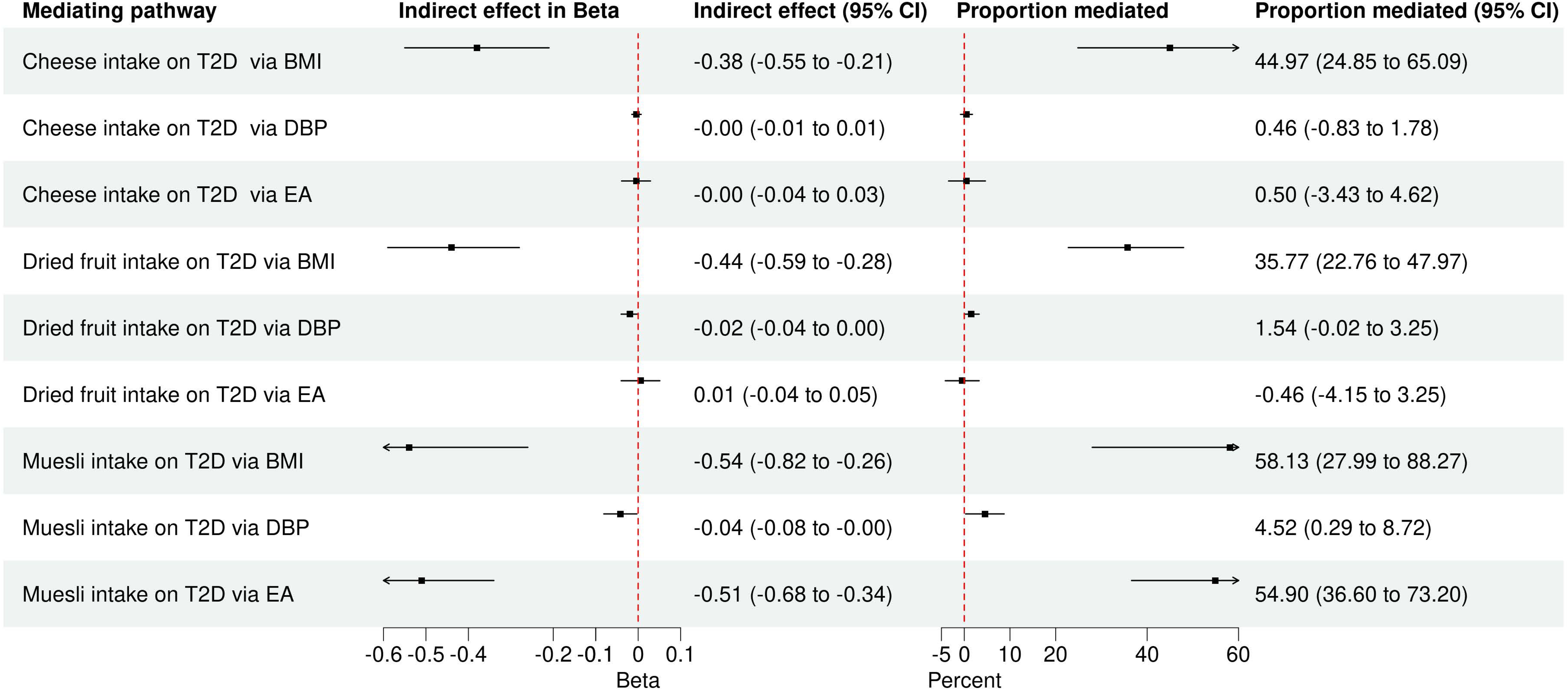
Forest plot for two-step MR mediation analysis. This figure is a forest plot for two-step MR mediation analysis that includes 95% confidence interval for estimated indirect effect and proportion mediated. It displays the indirect effects of 3 dietary preferences on T2D mediated individually by BMI, DBP and EA via RMediation package. Delta method (“asymptotic distribution”) was used to estimate the mediated effects by these 3 factors and calculate 95% confidence intervals. Consistent with MVMR, BMI was a major mediator for the 3 dietary associations with T2D and EA was additional mediator exclusively for the effect of muesli intake on T2D. Abbreviations – BMI, Body mass index; DBP, Diastolic blood pressure; EA, Educational attainment; MR, Mendelian Randomization; MVMR, Multivariable Mendelian Randomization; T2D, Type 2 diabetes mellitus.

Only BMI explained 44.97% (95% CI 24.85%, 65.09%) of the total effect of cheese intake on T2D, confirming the finding of MVMR that BMI is the only mediator for the protective effect of cheese intake on T2D. After adjustment for cheese intake, elevated BMI increased risk of T2D (IVW, β = 0.86; OR 2.36; 95% CI, 2.21 to 2.52; P = 1.20 x 10^-144^) but the associations of DBP and EA were not significant. The BMI-mediated effect constituted 35.77% (95% CI 22.76%, 47.97%) and 58.13% (95% CI 27.99%, 88.27%) of the total effects of dried fruit and muesli intake, respectively, on T2D.

The mediation analysis also helped clarify that DBP does not mediate the associations of dried fruit and muesli intake with T2D, because the DBP-mediate effects of dried fruit (1.54%, 95% CI -0.02%, 3.25%) and muesli intake (4.52%, 95% CI 0.29%, 8.72%) on T2D were minimal.

Consistent with MVMR, EA was another mediator for muesli intake-T2D association. Given that higher EA reduces the risk of T2D (IVW, β = -0.49; OR 0.61; 95% CI, 0.57 to 0.65; P = 1.20 x 10^-46^), EA explained 54.9% (95% CI 36.6%, 73.2%) of the total effect of muesli intake on T2D.

In agreement with MVMR, the mediation analysis demonstrated that BMI is a common factor that substantially mediates the associations of cheese, dried fruit and muesli intake with T2D.

## DISCUSSION

Type 2 diabetes mellitus (T2D) is a chronic disease that requires pharmacologic and lifestyle interventions for effective disease management. In particular, diet plays an important role in modulating the risk of T2D and influencing disease progression and onset of complications. Though past observational studies have shown that certain diets can promote good metabolic health or confer impaired glucose homeostasis [55,56], they cannot directly test causality.

Hence, we conducted Mendelian randomization (MR) study to examine genetic effects of various dietary preferences on T2D and 21 related cardiometabolic risk factors in order to determine which, if any, dietary exposures exerted causal effects on susceptibility to T2D or related traits. In univariable MR (UVMR), the most notable associations were between intake of cheese, dried fruits, muesli and spreads and T2D. These four inverse associations correspond to previously reported observational findings. Some prospective cohort studies found that total dairy consumption was associated with reduced T2D risk [57,58]. Cheese, a nutrient-dense dairy product with many bioactive compounds, has been observed to lower cardiovascular mortality and incidence of T2D [59]. Furthermore, the association between cheese intake and T2D could be partly explained by the interaction between lactase *LCT* gene and milk in the susceptibility of T2D [60,61]. High intake of whole grains, not refined grains, was shown to lower T2D risk [62]. The dose-response meta-analysis provided weak-to-moderate evidence that increasing servings of fiber-rich fruits and vegetables can help reduce incidence of T2D and related mortality [63,64]. Dietary fat is a low-glycemic source of energy and bioactive fatty acids that affect pancreatic beta cell metabolism, which is critical for glucose homeostasis [65]. Though total dietary fat consumption was not associated with T2D, some dietary fats, particularly regular dairy fats, have been shown to have benefits of reducing risk of T2D [66].

The follow-up multivariable MR experiments showed that socioeconomic and cardiometabolic risk factors play a mediating role in the observed associations. Importantly, body mass index (BMI) [67,68] was the factor that attenuated most of the dietary associations. These results reflect both the long-standing observation that obesity is a strong etiological risk factor for T2D, but also the close relationship between diet and adiposity. These results explain, in part, why some dietary preferences were shown to be associated with BMI including other adiposity related traits, such as WHR and MRI-based volumes of central, abdominal adipose tissues in UVMR. Surprisingly, none of the associations were affected by WHR, which is an alternative measure of adiposity. This observation might be due to difference in statistical power because GWAS data for BMI had a slightly larger sample size than for WHR. Alternatively, it might suggest that BMI and WHR are implicated in different pathways. In addition, we identified that EA was an additional mediating trait for muesli intake associations.

In the two-step MR mediation analysis, we were able to further interpret our initial observations from MVMR. The analysis further supported the specific diet preferences, particularly cheese intake, mediate susceptibility to T2D partly via BMI. It also concurred with MVMR that DBP was not a potential mediating factor in association between diet and T2D, aside from BMI and EA. Such observations suggest that adiposity and education are factors that diet interacts with to play an essential role in cardiometabolic health.

However, the study has limitations. Other than the three GWAS data sets for various dietary preference traits used for this study, there are no additional genetic data that are available in comparable sample size to replicate or further examine the findings. Also, the GWAS data for dietary preferences were specifically based on participants with European ancestry background that our results lack generalizability to other populations or ancestries. Because these dietary preferences are qualitatively measured via self-reported questionnaires from participants, they could be prone to bias. In the 2-step MR mediation analysis, the mediated (indirect) effect estimates should be taken with caution as the outcome of our interest was T2D, which is binary, that they were defined in noncollapsible odds ratio and therefore may not be accurate [69,70]. Lastly, because T2D is a multifactorial and heterogeneous disease, there could be unmeasured risk factors that were not tested in this study to further explain diet-T2D association.

Collectively, our findings show that diet is not an independent risk factor for T2D but dependent of established T2D risk factors, particularly obesity and low education level, to causally influence liability to T2D. These imply that diet plays a role in the maintenance of healthy weight and making food choices could be affected by education status and therefore is important for reducing T2D risk. The study also highlights that genetic data associated with diet should be integrated in tailoring effective dietary interventions for individuals at risk for poor metabolic health. Though further investigation is needed to confirm these findings, the current study provides positive genetic evidence that diet is one of the factors that contribute to susceptibility to T2D.

## Supporting information

STROBE-MR checklist

## ABBREVIATIONS

ALP: Alkaline phosphatase
ALT: Alanine transaminase
ASAT: Abdominal subcutaneous adipose tissue
AST: Aspartate aminotransferase
BMI: Body mass index
CORNFLAK: Corn flakes/frosties
DBP: Diastolic blood pressure
DRIEFRU: Dried fruit
EA: Educational attainment
FG: Fasting glucose adjusted for BMI
FI: Fasting insulin adjusted for BMI
GGT: Gamma-glutamyl transferase
GWAS: Genome-wide association study
HbA1c: Hemoglobin A1c
HDL: High-density lipoprotein
INFO: Information score (quality measurement of imputation)
IV: Instrumental variables
IVW: Inverse variance weighted
LDL: Low-density lipoprotein
LDSC: Linkage disequilibrium score regression
Liverfat: Liver fat content
Liveriron: Liver iron level
Livervol: Liver volume
MAF: Minor allele frequency
MR: Mendelian Randomization
MR-RAPS: Mendelian Randomization Robust Adjusted Profile Score
MVMR: Multivariable Mendelian Randomization
PAL: Highly palatable
Pancfat: Pancreas fat content
Panciron: Pancreas iron level
Pancvol: Pancreas volume
PHY: Physical activity
SAVCAL: Savory/ caloric
SBP: Systolic blood pressure
SED: Sedentary behavior at work
STR: Strong flavored
T2D: Type 2 diabetes mellitus
TG: Triglyceride
UVMR: Univariable Mendelian Randomization
VAT: Visceral adipose tissue
WHR: Wait-hip ratio
WHRadjBMI: Wait-hip ratio adjusted for BMI
WM: Weighted median

## ACKNOWLEDGEMENTS

Summary association data for type 2 diabetes obtained from the MVP project (phs001672) was obtained from dbGAP through data access request (approval #27398). BFV is grateful for support for the work from the NIH/NIDDK (DK126194). MDL is grateful for support for the work from the NIH/NHGRI (T32 HG000046).

## AUTHOR CONTRIBUTIONS

The authors’ responsibilities were as follows– MDL and BFV designed research and had primary responsibility for the final content; MDL conducted the analyses and wrote the manuscript; BFV supervised the project and contributed to the methodology used in the paper; MDL and BFV: interpreted the results, critically reviewed the manuscript, and approved the final manuscript.

## CONFLICT OF INTEREST

The authors report no conflicts of interest.

## SOFTWARE

Software Analyses were conducted in R software version 4.3.2, with the use of packages TwoSampleMR, MVMR [40,41,43] and RMediation [50].

## DATA AVAILABILITY

Data described in the manuscript, code book, and analytic code will be made publicly and freely available without restriction at https://github.com/mdayeon/Mendelian-randomization-on-diet-and-T2D.

## SUPPLEMENTARY FIGURE AND TABLE LEGENDS

**Supplementary Table 1:** GWAS data for dietary preferences used in MR.

**Supplementary Table 2:** GWAS data for T2D and 21 cardiometabolic risk factors used in MR.

**Supplementary Table 3:** Number of genetic instruments selected for each exposure.

**Supplementary Table 4:** *F*-statistics of the genetic instruments for exposures.

**Supplementary Table 5:** 17 significant associations (P < 5.99 x 10^-5^) in UVMR.

**Supplementary Table 6:** MR-Steiger test for the directions of the 17 associations in UVMR.

**Supplementary Table 7:** 17 Nominally significant (P < 1 x 10^-3^) associations in UVMR.

**Supplementary Table 8:** 7 significant associations (P < 2.94 x 10^-3^) in MR-RAPS.

**Supplementary Table 9:** MR-Steiger test for the directions of the 17 nominally significant associations in UVMR.

**Supplementary Table 10:** Re-assessment of spreads intake associations in MR-RAPS.

**Supplementary Table 11:** Single-mediator MVMR using 8 potential mediators.

**Supplementary Table 12:** Multiple-mediator MVMR (all-inclusive vs. non-BMI).

**Supplementary Table 13:** Single-mediator MVMR on 7 significant associations in MR-RAPS.

**Supplementary Table 14:** A table of genetic instruments for exposures, outcomes and potential mediators for UVMR and MVMR.

**Supplementary Table 15:** A table of genetic instruments for exposures, outcomes and potential mediators for MR-RAPS.

**Supplementary Table 16:** A summary table of UVMR on 835 exposure-outcome associations.

**Supplementary Table 17:** A table of genetic correlations between potential mediators and exposures.

**Supplementary Table 18:** A table of genetic correlations between potential mediators and outcomes.

**Supplementary Table 19:** 2-step MR mediation analysis on 3 dietary associations with T2D.

**Supplementary Table 20:** Indirect effect, proportion mediated and 95% confidence intervals via RMediation package.

**Supplementary Table 21:** Estimates of bias and Type 1 error rates due to sample overlap for the associations in UVMR and MR-RAPS.

**Supplementary Figure 1: Framework of MR**

Mendelian randomization employs genetic variants (e.g., SNPs) associated with an exposure as instrumental variables (IVs). Given that the selected IVs meet the 3 assumptions for IVs, the effect of an exposure on an outcome of interest is estimated.

**Supplementary Figure 2: Genetic instrument selection via PLINK**

The genetic instrument process was taken in 3 steps. First, variants that are rare (MAF < 0.01) or have low imputation quality (INFO < 0.5) were eliminated. Also, the datasets were screened for duplicate variants, if any. Second, a tool PLINK was used to perform clumping procedure and select genome-wide significant variants with low linkage disequilibrium. Last, only non-palindromic variants were taken into estimation of instrument strength using F-statistics formula to determine whether they are strong instruments that can be used in MR to test exposure-outcome associations.

**Supplementary Figure 3: Framework of Multivariable MR (MVMR)**

Multivariable MR (MVMR) assumes that genetic instruments selected are pleiotropic and therefore also associated with other traits (e.g., secondary exposure). The secondary exposure can be a mediator or confounder. It assesses the exposure-outcome association with inclusion of the secondary exposure. Whereas, univariable MR estimates the total effect of the exposure on the outcome, MVMR estimates the direct effect of exposure on outcome after adjusting for the effect of a potential confounder.

**Supplementary Figure 4: Forest plot for multiple-mediator MVMR**

Forest plot for multiple-mediator multivariable MR for comparison. The plot compares the summative effects of all mediators on the associations to that of all non-BMI mediators to determine whether BMI is the main driver for the causal effects of the dietary traits on T2D and cardiometabolic risk factors.

## Notes

### Competing Interest Statement

The authors have declared no competing interest.

### Funding Statement

This study did not receive any funding.

### Author Declarations

All the data used in this work were de-identified summary statistics data from genome-wide association studies. The publicly available GWAS summary-level datasets were made publicly available after approval by the institutional ethical committees of the different consortia. The most of the GWAS summary-level datasets used in the study were publicly available. The GWAS summary-level data for dietary composition (relative macronutrient intakes) was requested at https://www.thessgac.org/. The GWAS summary-level data for dietary habits and food liking traits were publicly available at https://www.kp4cd.org/dataset_downloads/t2d and The NHGRI-EBI GWAS Catalog website (https://www.ebi.ac.uk/gwas/publications/35585065), respectively. The GWAS summary-level data for type 2 diabetes mellitus was obtained from MVP study via request. The GWAS summary-level datasets for glycemic traits were publicly available at https://www.magicinvestigators.org/. The GWAS summary-level datasets for lipid traits were publicly available at http://csg.sph.umich.edu/willer/public/glgc-lipids2021/. The GWAS summary-level datasets for anthropometric traits were publicly available at https://portals.broadinstitute.org/collaboration/giant/index.php/GIANT_consortium_data_files. The GWAS summary-level datasets for abdominal organ MRI, liver enzymes and physical activity related traits were publicly available at The NHGRI-EBI GWAS Catalog website (https://www.ebi.ac.uk/gwas/publications/34128465, https://www.ebi.ac.uk/gwas/publications/33547301, https://www.ebi.ac.uk/gwas/publications/33972514, https://www.ebi.ac.uk/gwas/publications/36071172). The GWAS summary-level data for educational attainment was obtained via request at The NHGRI-EBI GWAS Catalog website (https://www.ebi.ac.uk/gwas/studies/GCST003676).

## REFERENCES

1. Ahlqvist E, Ahluwalia TS, Groop L. Genetics of type 2 diabetes. Clin Chem. 2011;57(2):241–54.

2. Ardisson Korat AV, Willett WC, Hu FB. Diet, lifestyle, and genetic risk factors for type 2 diabetes: a review from the Nurses’ Health Study, Nurses’ Health Study 2, and Health Professionals’ Follow-up Study. Curr Nutr Rep. 2014;3(4):345-354.

3. Zimmet PZ. Diabetes and its drivers: the largest epidemic in human history? Clin Diabetes Endocrinol. 2017;3:1.

4. Vujkovic M, Keaton JM, Lynch JA, Miller DR, Zhou J, Tcheandjieu C, et al. Discovery of 318 new risk loci for type 2 diabetes and related vascular outcomes among 1.4 million participants in a multi-ancestry meta-analysis. Nat Genet. 2020;52(7):680–691.

5. Mahajan A, Spracklen CN, Zhang W, Ng MCY, Petty LE, Kitajima H, et al. Multi-ancestry genetic study of type 2 diabetes highlights the power of diverse populations for discovery and translation. Nat Genet. 2022;54(5):560–572.

6. Cole JB, Florez JC. Genetics of diabetes mellitus and diabetes complications. Nat Rev Nephrol. 2020;16(7):377–390.

7. Parillo M, Riccardi G. Diet composition and the risk of type 2 diabetes: epidemiological and clinical evidence. Br J Nutr. 2004;92(1):7–19.

8. Guo Y, Huang Z, Sang D, Gao Q, Li Q. The Role of Nutrition in the Prevention and Intervention of Type 2 Diabetes. Front Bioeng Biotechnol. 2020;8:575442.

9. Koeth RA, Wang Z, Levison BS, Buffa JA, Org E, Sheehy BT, et al. Intestinal microbiota metabolism of L-carnitine, a nutrient in red meat, promotes atherosclerosis. Nat Med. 2013;19(5):576–85.

10. Laeger T, Henagan TM, Albarado DC, Redman LM, Bray GA, Noland RC, et al. FGF21 is an endocrine signal of protein restriction. J Clin Invest. 2014;124(9):3913–22.

11. Basu S, Yoffe P, Hills N, Lustig RH. The relationship of sugar to population-level diabetes prevalence: an econometric analysis of repeated cross-sectional data. PLoS One. 2013;8(2):e57873.

12. Luukkonen PK, Sädevirta S, Zhou Y, Kayser B, Ali A, Ahonen L, et al. Saturated Fat Is More Metabolically Harmful for the Human Liver Than Unsaturated Fat or Simple Sugars. Diabetes Care. 2018;41(8):1732–1739.

13. Hu FB, van Dam RM, Liu S. Diet and risk of Type II diabetes: the role of types of fat and carbohydrate. Diabetologia. 2001;44(7):805–17.

14. Tholin S, Rasmussen F, Tynelius P, Karlsson J. Genetic and environmental influences on eating behavior: the Swedish Young Male Twins Study. Am J Clin Nutr. 2005;81(3):564–9.

15. Hasselbalch AL, Heitmann BL, Kyvik KO, Sørensen TI. Studies of twins indicate that genetics influence dietary intake. J Nutr. 2008;138(12):2406–12.

16. Ortega Á, Berná G, Rojas A, Martín F, Soria B. Gene-Diet Interactions in Type 2 Diabetes: The Chicken and Egg Debate. Int J Mol Sci. 2017;18(6):1188.

17. Cole JB, Florez JC, Hirschhorn JN. Comprehensive genomic analysis of dietary habits in UK Biobank identifies hundreds of genetic associations. Nat Commun. 2020;11(1):1467.

18. Meddens SFW, de Vlaming R, Bowers P, Burik CAP, Linnér RK, Lee C, et al. Genomic analysis of diet composition finds novel loci and associations with health and lifestyle. Mol Psychiatry. 2021;26(6):2056–2069.

19. May-Wilson S, Matoba N, Wade KH, Hottenga JJ, Concas MP, Mangino M, et al. Large-scale GWAS of food liking reveals genetic determinants and genetic correlations with distinct neurophysiological traits. Nat Commun. 2022;13(1):2743.

20. Burgess S, Small DS, Thompson SG. A review of instrumental variable estimators for Mendelian randomization. Stat Methods Med Res. 2017;26(5):2333–2355.

21. Sanderson E, Glymour MM, Holmes MV, Kang H, Morrison J, Munafò MR, et al. Mendelian randomization. Nat Rev Methods Primers. 2022;2:6.

22. Yuan S, Larsson SC. An atlas on risk factors for type 2 diabetes: a wide-angled Mendelian randomisation study. Diabetologia. 2020;63(11):2359–2371.

23. Ference BA, Robinson JG, Brook RD, Catapano AL, Chapman MJ, Neff DR, et al. Variation in PCSK9 and HMGCR and Risk of Cardiovascular Disease and Diabetes. N Engl J Med. 2016;375(22):2144–2153.

24. Holmes MV, Dale CE, Zuccolo L, Silverwood RJ, Guo Y, Ye Z, et al. Association between alcohol and cardiovascular disease: Mendelian randomisation analysis based on individual participant data. BMJ. 2014;349:g4164.

25. Larsson SC, Carter P, Kar S, Vithayathil M, Mason AM, Michaëlsson K, et al. Smoking, alcohol consumption, and cancer: A mendelian randomisation study in UK Biobank and international genetic consortia participants. PLoS Med. 2020;17(7):e1003178.

26. Cohen JC, Boerwinkle E, Mosley TH Jr, Hobbs HH. Sequence variations in PCSK9, low LDL, and protection against coronary heart disease. N Engl J Med. 2006;354(12):1264–72.

27. Yao S, Zhang M, Dong SS, Wang JH, Zhang K, Guo J, et al. Bidirectional two-sample Mendelian randomization analysis identifies causal associations between relative carbohydrate intake and depression. Nat Hum Behav. 2022;6(11):1569–1576.

28. Pulit SL, Stoneman C, Morris AP, Wood AR, Glastonbury CA, Tyrrell J, et al. Meta-analysis of genome-wide association studies for body fat distribution in 694 649 individuals of European ancestry. Hum Mol Genet. 2019;28(1):166–174.

29. Chen J, Spracklen CN, Marenne G, Varshney A, Corbin LJ, Luan J, et al. The trans-ancestral genomic architecture of glycemic traits. Nat Genet. 2021;53(6):840–860.

30. Graham SE, Clarke SL, Wu KH, Kanoni S, Zajac GJM, Ramdas S, et al. The power of genetic diversity in genome-wide association studies of lipids. Nature. 202;600(7890):675-679.

31. Liu Y, Basty N, Whitcher B, Bell JD, Sorokin EP, van Bruggen N, et al. Genetic architecture of 11 organ traits derived from abdominal MRI using deep learning. Elife. 2021;10:e65554.

32. Pazoki R, Vujkovic M, Elliott J, Evangelou E, Gill D, Ghanbari M, et al. Genetic analysis in European ancestry individuals identifies 517 loci associated with liver enzymes. Nat Commun. 2021;12(1):2579.

33. Chen VL, Du X, Chen Y, Kuppa A, Handelman SK, Vohnoutka RB, et al. Genome-wide association study of serum liver enzymes implicates diverse metabolic and liver pathology. Nat Commun. 2021;12(1):816.

34. Purcell S, Neale B, Todd-Brown K, Thomas L, Ferreira MA, Bender D, et al. PLINK: a tool set for whole-genome association and population-based linkage analyses. Am J Hum Genet. 2007;81(3):559–75.

35. Burgess S, Davey Smith G, Davies NM, Dudbridge F, Gill D, Glymour MM, et al. Guidelines for performing Mendelian randomization investigations: update for summer 2023. Wellcome Open Res. 2023;4:186.

36. Brion MJ, Shakhbazov K, Visscher PM. Calculating statistical power in Mendelian randomization studies. Int J Epidemiol. 2013;42(5):1497–501.

37. Burgess S, Thompson SG. Bias in causal estimates from Mendelian randomization studies with weak instruments. Stat Med. 2011;30(11):1312–23.

38. Zhao Q, Wang J, Hemani G, Bowden J, Small DS. Statistical inference in two-sample summary-data Mendelian randomization using robust adjusted profile score. Ann Statist. 2020;48(3):1742–1769.

39. Burgess S, Davies NM, Thompson SG. Bias due to participant overlap in two-sample Mendelian randomization. Genet Epidemiol. 2016;40(7):597–608.

40. Hemani G, Zheng J, Elsworth B, Wade KH, Haberland V, Baird D, et al. The MR-Base platform supports systematic causal inference across the human phenome. Elife. 2018;7:e34408.

41. Hemani G, Tilling K, Davey Smith G. Orienting the causal relationship between imprecisely measured traits using GWAS summary data. PLoS Genet. 2017;13(11):e1007081.

42. Slob EAW, Burgess S. A comparison of robust Mendelian randomization methods using summary data. Genet Epidemiol. 2020;44(4):313–329.

43. Sanderson E, Spiller W, Bowden J. Testing and correcting for weak and pleiotropic instruments in two-sample multivariable Mendelian randomization. Stat Med. 2021;40(25):5434–5452.

44. Bulik-Sullivan BK, Loh PR, Finucane HK, Ripke S, Yang J, Schizophrenia Working Group of the Psychiatric Genomics Consortium; et al. LD Score regression distinguishes confounding from polygenicity in genome-wide association studies. Nat Genet. 2015;47(3):291–5.

45. Papaioannou TG, Oikonomou E, Lazaros G, Christoforatou E, Vogiatzi G, Tsalamandris S, et al. Arterial stiffness and subclinical aortic damage of reclassified subjects as stage 1 hypertension according to the new 2017 ACC/AHA blood pressure guidelines. Vasa. 2019;48(3):236–243.

46. Wang Z, Emmerich A, Pillon NJ, Moore T, Hemerich D, Cornelis MC, et al. Genome-wide association analyses of physical activity and sedentary behavior provide insights into underlying mechanisms and roles in disease prevention. Nat Genet. 2022;54(9):1332–1344.

47. Okbay A, Wu Y, Wang N, Jayashankar H, Bennett M, Nehzati SM, et al. Polygenic prediction of educational attainment within and between families from genome-wide association analyses in 3 million individuals. Nat Genet. 2022;54(4):437–449.

48. Carter AR, Sanderson E, Hammerton G, Richmond RC, Davey Smith G, Heron J, et al. Mendelian randomisation for mediation analysis: current methods and challenges for implementation. Eur J Epidemiol. 2021;36(5):465-478.

49. Burgess S, Daniel RM, Butterworth AS, Thompson SG, EPIC-InterAct Consortium. Network Mendelian randomization: using genetic variants as instrumental variables to investigate mediation in causal pathways. Int J Epidemiol. 2015;44(2):484–95.

50. Tofighi D, MacKinnon DP. RMediation: an R package for mediation analysis confidence intervals. Behav Res Methods. 2011;43(3):692–700.

51. Thompson JR, Minelli C, Del Greco M F. Mendelian Randomization using Public Data from Genetic Consortia. Int J Biostat. 2016;12(2):/j/ijb.2016.12.issue-2/ijb-2015-0074/ijb-2015-0074.xml.

52. Carter AR, Gill D, Davies NM, Taylor AE, Tillmann T, Vaucher J, et al. Understanding the consequences of education inequality on cardiovascular disease: mendelian randomisation study. BMJ. 2019;365:l1855.

53. Sung KC, Seo MH, Rhee EJ, Wilson AM. Elevated fasting insulin predicts the future incidence of metabolic syndrome: a 5-year follow-up study. Cardiovasc Diabetol. 2011;10:108.

54. ter Horst KW, Gilijamse PW, Koopman KE, de Weijer BA, Brands M, Kootte RS, et al. Insulin resistance in obesity can be reliably identified from fasting plasma insulin. Int J Obes (Lond). 2015;39(12):1703–9.

55. Martín-Peláez S, Fito M, Castaner O. Mediterranean Diet Effects on Type 2 Diabetes Prevention, Disease Progression, and Related Mechanisms. A Review. Nutrients. 2020;12(8):2236.

56. Neuenschwander M, Ballon A, Weber KS, Norat T, Aune D, Schwingshackl L, et al. Role of diet in type 2 diabetes incidence: umbrella review of meta-analyses of prospective observational studies. BMJ. 2019;366:l2368.

57. Choi HK, Willett WC, Stampfer MJ, Rimm E, Hu FB. Dairy consumption and risk of type 2 diabetes mellitus in men: a prospective study. Arch Intern Med. 2005;165(9):997–1003.

58. Pittas AG, Dawson-Hughes B, Li T, Van Dam RM, Willett WC, Manson JE, et al. Vitamin D and calcium intake in relation to type 2 diabetes in women. Diabetes Care. 2006;29(3):650–6.

59. Zhang M, Dong X, Huang Z, Li X, Zhao Y, Wang Y, et al. Cheese consumption and multiple health outcomes: an umbrella review and updated meta-analysis of prospective studies. Adv Nutr. 2023;14(5):1170–1186.

60. Luo K, Chen GC, Zhang Y, Moon JY, Xing J, Peters BA, et al. Variant of the lactase LCT gene explains association between milk intake and incident type 2 diabetes. Nat Metab. 2024;6(1):169–186.

61. Littleton SH, Grant SFA. Metabolic links among milk, genes and gut. Nat Metab. 2024;6(1):12–13.

62. Aune D, Norat T, Romundstad P, Vatten LJ. Whole grain and refined grain consumption and the risk of type 2 diabetes: a systematic review and dose-response meta-analysis of cohort studies. Eur J Epidemiol. 2013;28(11):845–58.

63. Wang DD, Li Y, Bhupathiraju SN, Rosner BA, Sun Q, Giovannucci EL, et al. Fruit and Vegetable Intake and Mortality: Results From 2 Prospective Cohort Studies of US Men and Women and a Meta-Analysis of 26 Cohort Studies. Circulation. 2021;143(17):1642–1654.

64. Li M, Fan Y, Zhang X, Hou W, Tang Z. Fruit and vegetable intake and risk of type 2 diabetes mellitus: meta-analysis of prospective cohort studies. BMJ Open. 2014;4(11):e005497.

65. Acosta-Montaño P, García-González V. Effects of Dietary Fatty Acids in Pancreatic Beta Cell Metabolism, Implications in Homeostasis. Nutrients. 2018;10(4):393.

66. Rice Bradley BH. Dietary Fat and Risk for Type 2 Diabetes: a Review of Recent Research. Curr Nutr Rep. 2018;7(4):214–226.

67. Narayan KM, Boyle JP, Thompson TJ, Gregg EW, Williamson DF. Effect of BMI on lifetime risk for diabetes in the U.S. Diabetes Care. 2007;30(6):1562–6.

68. Gray N, Picone G, Sloan F, Yashkin A. Relation between BMI and diabetes mellitus and its complications among US older adults. South Med J. 2015;108(1):29–36.

69. Pang M, Kaufman JS, Platt RW. Studying noncollapsibility of the odds ratio with marginal structural and logistic regression models. Stat Methods Med Res. 2016;25(5):1925–1937.

70. Vanderweele TJ, Vansteelandt S. Odds ratios for mediation analysis for a dichotomous outcome. Am J Epidemiol. 2010;172(12):1339–48.

