## Supplementary Figures for "Dietary preference and susceptibility to type 2 diabetes mellitus: a wide-angle Mendelian randomization study"

#### Supplementary Figure 1: MR framework for UVMR

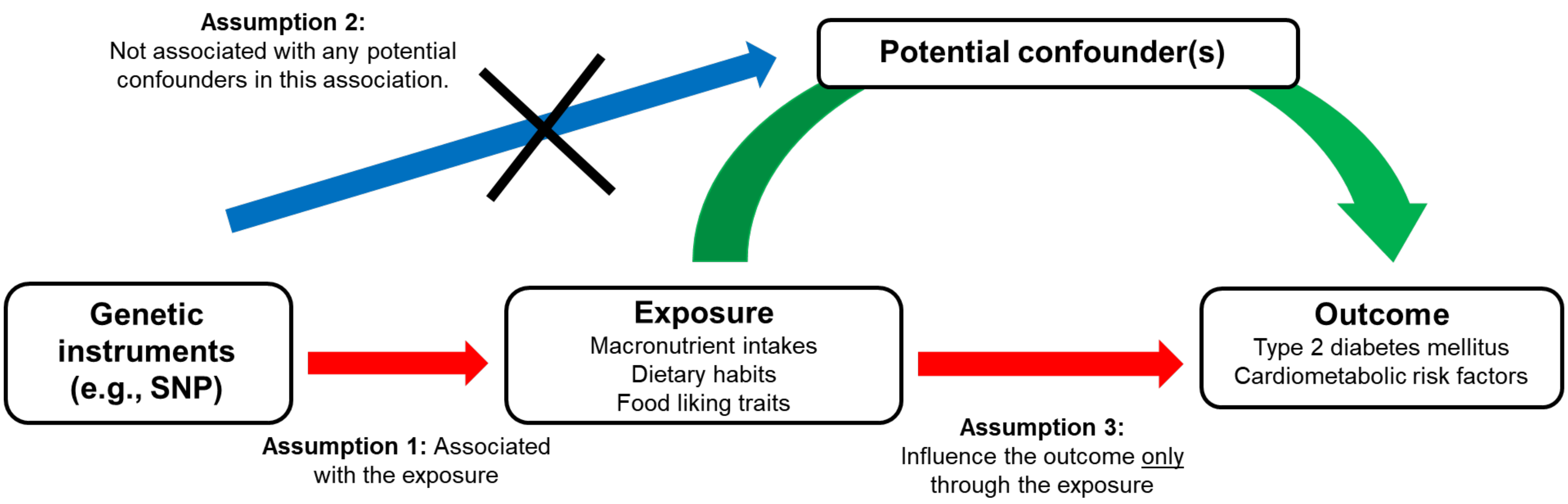


**Supplementary Figure 1.** Mendelian randomization employs genetic variants (e.g., SNPs) associated with an exposure as instrumental variables (IVs). Given that the selected IVs meet the 3 assumptions (as shown in the above) for IVs, the effect of an exposure on an outcome of interest is estimated.

### Supplementary Figure 2: Clumping procedure / genetic instrument selection

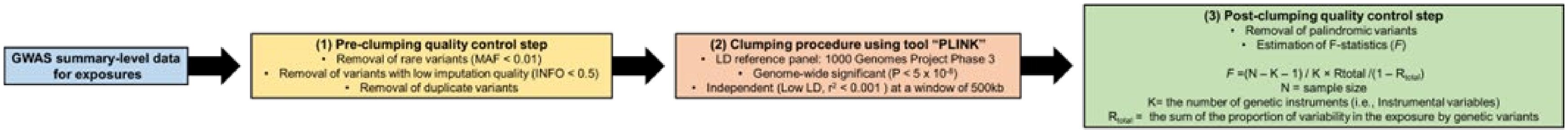


**Supplementary Figure 2.** After GWAS datasets for the exposures of interest (dietary preferences) were obtained, the genetic instrument process was taken in 3 steps. First, variants that are rare (MAF < 0.01) or have low imputation quality (INFO < 0.5) were eliminated. Also, the datasets were screened for duplicate variants, if any. Second, a tool PLINK was used to perform clumping procedure and select genome-wide significant variants with low linkage disequilibrium. Last, only non-palindromic variants were taken into estimation of instrument strength using *F*-statistics formula to determine whether they are strong instruments that can be used in MR to test exposure:outcome associations.

**Supplementary Figure 3: MR framework for MVMR**

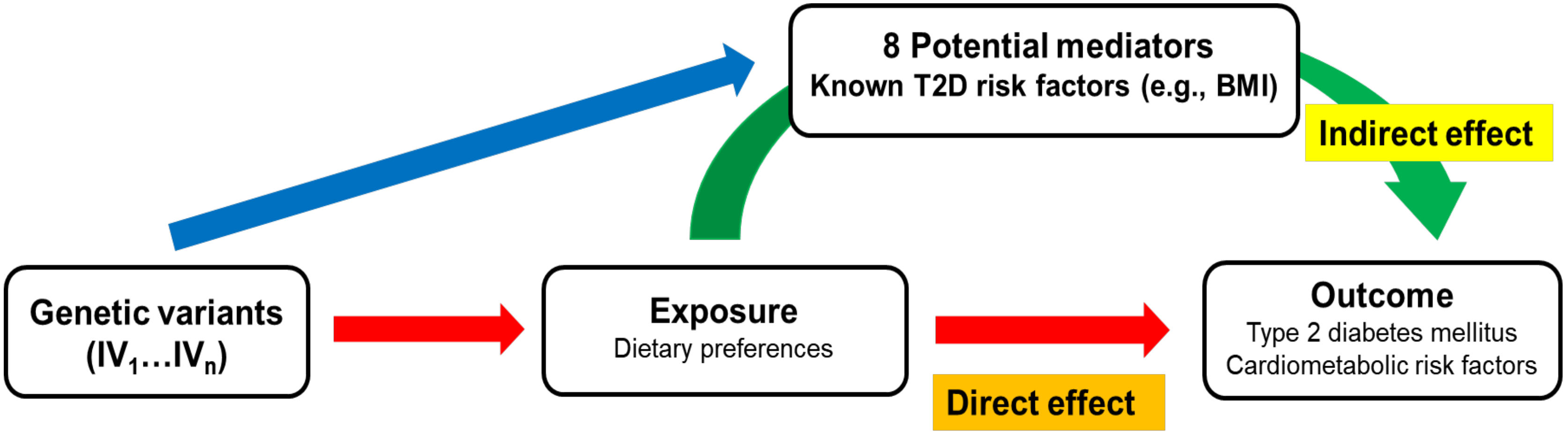


**Supplementary Figure 3**. Multivariable MR (MVMR) assumes that genetic instruments selected are pleiotropic and therefore also associated with other trait (e.g., secondary exposure). The secondary exposure can be a mediator or confounder. It assesses the exposure:outcome association with inclusion of the secondary exposure. Whereas, univariable MR estimates the total effect of the exposure on the outcome, MVMR estimates the direct effect of exposure on outcome after adjusting for the effect of a potential confounder.

**Supplementary Figure 4:** Forest plot for multiple-mediator MVMR: non-BMI MVMR vs. all-inclusive, BMI-only MVMR using BMI vs. UVMR

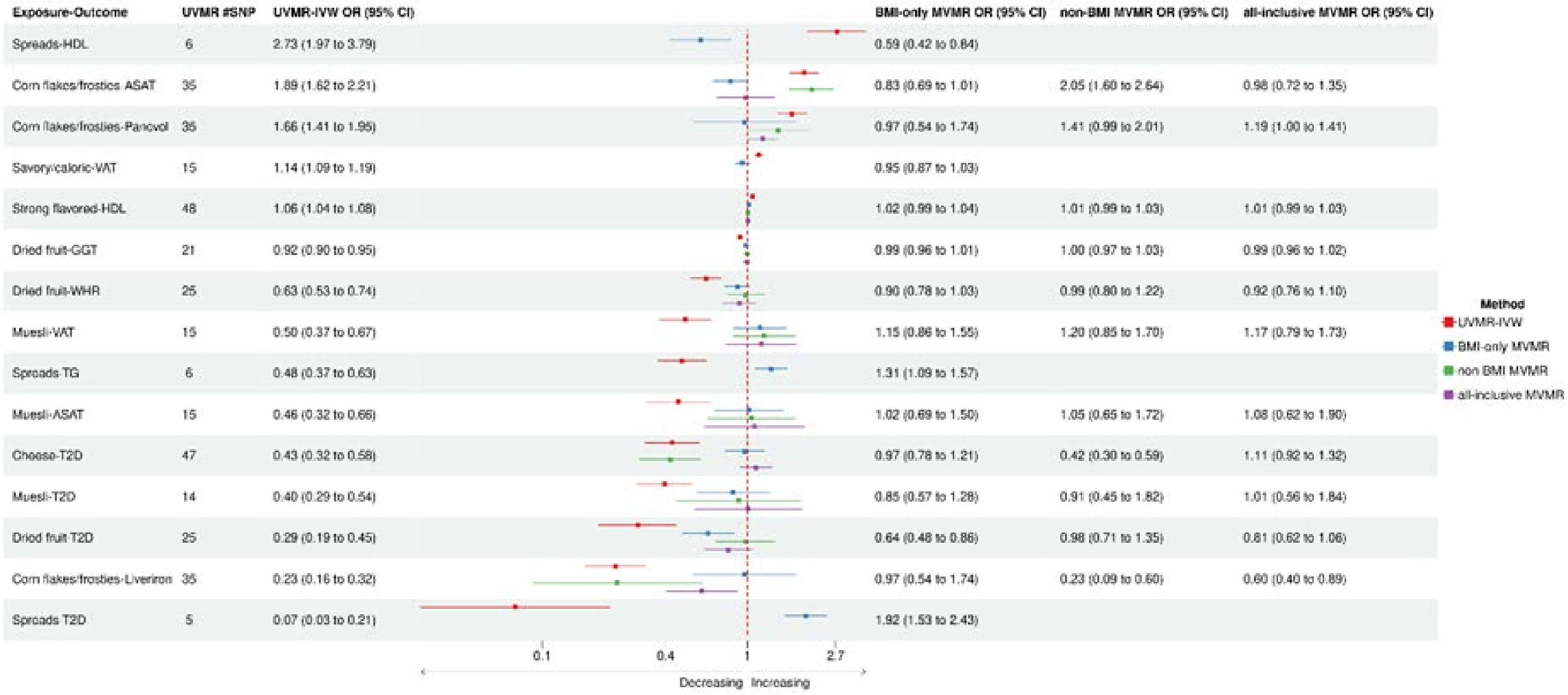


**Supplementary Figure 4.** Forest plot for multiple-mediator multivariable MR for comparison. The plot compares the summative effects of all mediators on the associations to that of all non-BMI mediators to determine whether BMI is the main driver for the causal effects of the dietary traits on T2D and cardiometabolic risk factors.
