## Supplementary material for "Dietary preference and susceptibility to type 2 diabetes mellitus: a wide-angle Mendelian randomization study": STROBE-MR checklist

**STROBE-MR checklist of recommended items to address in reports of Mendelian randomization studies**[1 2]

| Item No. | Section | | Checklist item | Page No. | Relevant text from manuscript |
|---|---|---|---|---|---|
| 1 | **TITLE and ABSTRACT** | | Indicate Mendelian randomization (MR) as the study's design in the title and/or the abstract if that is a main purpose of the study | | Complete (as shown in the Title page - 1st page) |
| | **INTRODUCTION** | | | | |
| 2 | **Background** | | Explain the scientific background and rationale for the reported study. What is the exposure? Is a potential causal relationship between exposure and outcome plausible? Justify why MR is a helpful method to address the study question | | Complete; In the last paragraph of Introduction, we explain the rationale for choosing MR for the study and specify the exposure traits of interest; In the *MR methods* of Methods, we further describe how MR works. |
| 3 | **Objectives** | | State specific objectives clearly, including pre-specified causal hypotheses (if any). State that MR is a method that, under specific assumptions, intends to estimate causal effects | | Complete; In Introduction, we state our primary objective of assessing causality in diet and T2D via MR as to address limitations of observational studies; In Methods, we also describe the assumptions for MR analysis to estimate causal effects |
| | **METHODS** | | | | |
| 4 | **Study design and data sources** | | Present key elements of the study design early in the article. Consider including a table listing sources of data for all phases of the study. For each data source contributing to the analysis, describe the following: | | Complete; See Figure 1 - workflow |
| | | a) | Setting: Describe the study design and the underlying population, if possible. Describe the setting, locations, and relevant dates, including periods of recruitment, exposure, follow-up, and data collection, when available. | | Complete; In Methods, the exposure and outcome traits tested are specified with description of where the data for those traits were obtained from; Also, Supp Tables 1-2 contain details of data sources for those traits. |
| | | b) | Participants: Give the eligibility criteria, and the sources and methods of selection of participants. Report the sample size, and whether any power or sample size calculations were carried out prior to the main analysis | | Complete; In Methods, *Acquisition of dietary trait summary genetic data* and *Acquisition of T2D and related traits summary genetic data* describe the cohorts and their ancestry and sample sizes of the exposure and outcome datasets selected; Also, Supp Tables 1-2 contain further details. |
| | | c) | Describe measurement, quality control and selection of genetic variants | | Complete; In *GWAS data processing and quality control* & *Selection of variants for genetic instruments for dietary traits* of Methods, we describe clumping procedure via PLINK and the filters applied for quality control for genetic |

| | | | | |
|---|---|---|---|---|
| | | | | instrument selection. We also explain that F-statistics was calculated to determine the strengths of the selected genetic instruments; See Supp. Figures 1-2 and 4. |
| | | d) | For each exposure, outcome, and other relevant variables, describe methods of assessment and diagnostic criteria for diseases | Complete; In Methods, *GWAS data processing and quality control* & *Selection of variants for genetic instruments for dietary traits* explain how F-statistics was calculated for genetic instruments selected for exposure traits. Aside from T2D, we selected additional outcome traits that are known, established cardiometabolic risk factors for T2D. |
| | | e) | Provide details of ethics committee approval and participant informed consent, if relevant | n/a |
| 5 | **Assumptions** | | Explicitly state the three core IV assumptions for the main analysis (relevance, independence and exclusion restriction) as well assumptions for any additional or sensitivity analysis | Complete; In Methods, *Mendelian randomization methods* explains the 3 core assumptions for IVs (genetic instruments) when using MR analysis to estimate causal effects. Each assumption was named accordingly. |
| 6 | **Statistical methods: main analysis** | | Describe statistical methods and statistics used | Complete; In Methods, *Mendelian randomization methods* describes the 3 standard methods used for univariable analysis. Also, *Analysis using MR-RAPS (Robust Adjusted Profile Score)* & *Multivariable MR experimental methods* describe the process of how MR-RAPS and multivariable analyses were conducted. |
| | | a) | Describe how quantitative variables were handled in the analyses (i.e., scale, units, model) | Complete; Since dietary exposure traits have different measurement units/scale (e.g., kcal, cups/day, pieces/day, times/week - categorical, ordinal), we applied “amount of intake” for each dietary exposure-outcome association tested. In Methods, Acquisition of dietary trait summary genetic data explains the scales/units used for dietary exposure traits in GWAS; See Supp. Table 1. |
| | | b) | Describe how genetic variants were handled in the analyses and, if applicable, how their weights were selected | Complete; weights for the dietary exposure traits were taken from the relevant GWAS for those exposures (selecting the lead variant tag and effect size), described in Methods. Weights for multivariable analyses also derive from their associated GWAS as described in Methods. |

| | | | | |
|---|---|---|---|---|
| | | c) | Describe the MR estimator (e.g. two-stage least squares, Wald ratio) and related statistics. Detail the included covariates and, in case of two-sample MR, whether the same covariate set was used for adjustment in the two samples | Complete; In Methods, *Mendelian randomization methods* describes 3 standard methods used in two-sample univariable analysis; In Methods, *MR-RAPS analysis* and *Multivariable MR experiments* describe additional analyses followed by univariable analysis. Also, the covariates used to conduct association analysis and generate summary association data for exposures, outcomes and mediators were listed in *Acquisition of dietary trait summary genetic data* in Methods and Supp Table 2. |
| | | d) | Explain how missing data were addressed | Completed; Because some exposure traits had insufficient genetic instruments, we used PLINK to obtain proxy genetic variants for multivariable analysis. We describe the process in *Selection of variants for genetic instruments for dietary traits* in Methods. |
| | | e) | If applicable, indicate how multiple testing was addressed | Complete; In Methods, *Mendelian randomization methods* describes that we used Bonferroni correction on threshold p-value to address multiple testing burden. |
| 7 | **Assessment of assumptions** | | Describe any methods or prior knowledge used to assess the assumptions or justify their validity | No specific priors or methods were applied to assess assumptions or justify validity. We apply methods robust to violation of some assumptions as described in text (e.g., median-weighted, MVMR) |
| 8 | **Sensitivity analyses and additional analyses** | | Describe any sensitivity analyses or additional analyses performed (e.g. comparison of effect estimates from different approaches, independent replication, bias analytic techniques, validation of instruments, simulations) | Complete; In Methods, we describe all analyses including two-step mediation analysis performed for the study. |
| 9 | **Software and pre-registration** | | | |
| | | a) | Name statistical software and package(s), including version and settings used | Complete; In Methods, we indicated the versions of PLINK, TwoSampleMR R package, MVMR R package and RMediation R package. |
| | | b) | State whether the study protocol and details were pre-registered (as well as when and where) | n/a |
| | **RESULTS** | | | |

| | | | | |
|---|---|---|---|---|
| 10 | **Descriptive data** | | | |
| | | a) | Report the numbers of individuals at each stage of included studies and reasons for exclusion. Consider use of a flow diagram | Complete; In the first paragraph of Results, we indicate that we tested 38 dietary exposures that had minimum 5 genetic instruments in univariable analysis. We explained that, of 835 associations, the ones where <5 instruments were used were eliminated from consideration. See Figure 1. |
| | | b) | Report summary statistics for phenotypic exposure(s), outcome(s), and other relevant variables (e.g. means, SDs, proportions) | Summary stats for phenotypic exposures are available from primary (public) data from which these exposures derive; outcomes are more complex as they derive from meta-analysis, but cohort data are routinely provided in the supplementary material of those works. |
| | | c) | If the data sources include meta-analyses of previous studies, provide the assessments of heterogeneity across these studies | This was not evaluated. |
| | | d) | For two-sample MR:<br>i. Provide justification of the similarity of the genetic variant-exposure associations between the exposure and outcome samples<br>ii. Provide information on the number of individuals who overlap between the exposure and outcome studies | Sample overlap exists between the dietary exposure traits and some outcome traits; however, the strength of instruments suggests that the observed overlap will only contribute small amount of bias (see PMID: 27625185) |
| 11 | **Main results** | | | |
| | | a) | Report the associations between genetic variant and exposure, and between genetic variant and outcome, preferably on an interpretable scale | Complete; Supp. Table 14 lists the selected genetic variants for exposure and outcome traits. |
| | | b) | Report MR estimates of the relationship between exposure and outcome, and the measures of uncertainty from the MR analysis, on an interpretable scale, such as odds ratio or relative risk per SD difference | Complete; In Results, we present 17 associations and their estimates from standard univariable MR and 7 association from MR-RAPS analysis. See Figure 2, Supp. Table 5, Figure 3, Supp. Table 8. |
| | | c) | If relevant, consider translating estimates of relative risk into absolute risk for a meaningful time period | n/a |
| | | d) | Consider plots to visualize results (e.g. forest plot, scatterplot of associations between genetic variants and outcome versus between genetic variants and exposure) | Complete; forest plots were generated to visualize all MR analyses including mediation analysis. In Results. See Fig 3-5, 6a-b, 7, and Supp Fig 4, , |
| 12 | **Assessment of assumptions** | | | |

| | | | | |
|---|---|---|---|---|
| | | a) | Report the assessment of the validity of the assumptions | We did not perform additional formal assessments of validity of MR assumptions beyond investigation and use of varied MR methods reported here. |
| | | b) | Report any additional statistics (e.g., assessments of heterogeneity across genetic variants, such as $I^2$, Q statistic or E-value) | n/a |
| 13 | **Sensitivity analyses and additional analyses** | | | |
| | | a) | Report any sensitivity analyses to assess the robustness of the main results to violations of the assumptions | Complete; As described in Methods and Results, methods Weighted median and Egger regression were employed in addition to inverse variance weighted for univariable analysis. |
| | | b) | Report results from other sensitivity analyses or additional analyses | Complete; See Results, Supp Tables 5-7, 10-13, 19, 20 |
| | | c) | Report any assessment of direction of causal relationship (e.g., bidirectional MR) | Complete; As described in *Mendelian randomization methods* of Methods, MR-Steiger tests were conducted on the observed associations for the correct directions of causality; See Supp Tables 6 and 9. |
| | | d) | When relevant, report and compare with estimates from non-MR analyses | Complete; As described in *Mediation analysis* in Methods and Results, two-step mediation analysis were conducted to estimate indirect effects to further evaluate and compare with multivariable analysis results. |
| | | e) | Consider additional plots to visualize results (e.g., leave-one-out analyses) | n/a |
| | **DISCUSSION** | | | |
| 14 | **Key results** | | Summarize key results with reference to study objectives | Complete; See paragraphs 1-4 in Discussion |
| 15 | **Limitations** | | Discuss limitations of the study, taking into account the validity of the IV assumptions, other sources of potential bias, and imprecision. Discuss both direction and magnitude of any potential bias and any efforts to address them | Complete; See paragraph 5 |
| 16 | **Interpretation** | | | |
| | | a) | Meaning: Give a cautious overall interpretation of results in the context of their limitations and in comparison with other studies | Complete; Throughout Discussion |

| | | | |
|---|---|---|---|
| | | b) Mechanism: Discuss underlying biological mechanisms that could drive a potential causal relationship between the investigated exposure and the outcome, and whether the gene-environment equivalence assumption is reasonable. Use causal language carefully, clarifying that IV estimates may provide causal effects only under certain assumptions | Complete; See paragraphs 3,4,6 |
| | | c) Clinical relevance: Discuss whether the results have clinical or public policy relevance, and to what extent they inform effect sizes of possible interventions | Complete; See paragraph 6 in Discussion |
| 17 | **Generalizability** | Discuss the generalizability of the study results (a) to other populations, (b) across other exposure periods/timings, and (c) across other levels of exposure | n/a |
| | **OTHER INFORMATION** | | |
| 18 | **Funding** | Describe sources of funding and the role of funders in the present study and, if applicable, sources of funding for the databases and original study or studies on which the present study is based | n/a |
| 19 | **Data and data sharing** | Provide the data used to perform all analyses or report where and how the data can be accessed, and reference these sources in the article. Provide the statistical code needed to reproduce the results in the article, or report whether the code is publicly accessible and if so, where | We followed the journal guidelines for data and data sharing requirements. The statistical code is publicly accessible via GitHub repository |
| 20 | **Conflicts of Interest** | All authors should declare all potential conflicts of interest | We completed conflicts of interest (if any) statement. |

This checklist is copyrighted by the Equator Network under the Creative Commons Attribution 3.0 Unported (CC BY 3.0) license.

1. Skrivankova VW, Richmond RC, Woolf BAR, Yarmolinsky J, Davies NM, Swanson SA, et al. Strengthening the Reporting of Observational Studies in Epidemiology using Mendelian Randomization (STROBE-MR) Statement. JAMA. 2021;under review.

2. Skrivankova VW, Richmond RC, Woolf BAR, Davies NM, Swanson SA, VanderWeele TJ, et al. Strengthening the Reporting of Observational Studies in Epidemiology using Mendelian Randomisation (STROBE-MR): Explanation and Elaboration. BMJ. 2021;375:n2233.
